# B cell-extrinsic and intrinsic factors linked to early immune repletion after anti-CD20 therapy in patients with Multiple Sclerosis of African Ancestry

**DOI:** 10.1101/2025.03.05.25321882

**Authors:** Gregg J. Silverman, Abhimanyu N. Amarnani, Arnaldo A. Armini, Angie Kim, Hannah Kopinsky, David Fenyo, Ilya Kister

## Abstract

In prior investigations, we identified patients of African ancestry (AA) with Multiple Sclerosis (MS) who displayed more rapid B-cell repopulation after treatment with anti-CD20 monoclonal antibodies. In this study, we explored immunologic, serum drug monitoring, and genetic factors that may contribute to a faster rate of B-cell repletion in AA patients with MS, with comparisons to those with usual repletion patterns after semi-annual infusions. Our assessment of extrinsic factors revealed an unexpected prevalence of anti-drug antibodies against ocrelizumab with concurrent undetectable serum drug levels in this patient subset. Considering intrinsic factors, a separate set of ER patients of African descent, without anti-drug antibodies, showed a significant overrepresentation of genetic polymorphisms, that included single nucleotide polymorphisms (SNPs) that map to genes for the B cell survival factor (BAFF) and antibody-dependent cytotoxicity, as well as pathways involved in the inflammatory response, leukocyte activation, and B cell differentiation. Larger studies are now needed to determine whether these genetic factors in AA MS patients, associated with early repletion, may lead to impaired therapeutic benefits and clinical progression, and the findings could be used to guide future personalized therapeutic strategies.

## Introduction

Multiple sclerosis (MS), a chronic demyelinating disease that affects the central nervous system (CNS), was traditionally viewed as largely being a ‘T-cell mediated’ disorder, in part because of the predominance of T cells in demyelinating plaques. However, it is now recognized that MS-related pathological processes involve interactions between several immune cell types, with increasing evidence that antibody-independent functions of B cells play key roles in mediating disease activity^1-5^. The potential roles of B-cells in MS have been highlighted, in part, by their increased representation in CSF of MS patients^4,5^ and the efficacy of B-cell-depleting therapies in suppressing relapses and new MRI lesions in MS^6,7^. Treatments involve regular antibody injections or infusions, resulting in B cell depletion in the bloodstream, with variability in the level of tissue B cell depletion and the time to B cell repopulation of the circulation.

MS affects individuals from various races and ethnicities^8^. Recent studies indicate that the incidence of MS in the US was highest among women of African ancestry (AA), relative to other racial-ethnic groups^9^, yet MS patients of African descent remain understudied^10^. In our large and highly diverse urban MS cohort, we found that African ancestry was a predictor of faster B-cell repletion among anti-CD20 therapy-treated patients^11^. To investigate mechanisms responsible for early repletion, we have performed a range of cellular, serum antibody, protein assays, and genetic analyses in AA MS patients with and without early repletion. The clinical consequences of early B cell repletion were also assessed.

## Materials and Methods

### Clinical and demographic characteristics and selection criteria

This study was approved by NYU Langone IRB. All patients signed informed consent. Eligible patients self-identified as AA and were receiving their neurologic care at the NYU MS Care Center. All but three patients met the 2017 revised McDonald Criteria^12^ for relapsing-remitting MS, and these patients met the criteria for Neuromyelitis optica spectrum disorder (NMOSD). All participants were receiving infusions of anti-CD20 antibody therapy (either rituximab or ocrelizumab) at the time of enrollment. All participants were recruited between July 2022 and May 2023.

Demographic, clinical characteristics, treatment history as well as details on relapses and MRI activity while on anti-CD20 therapy were abstracted from the medical record by two neurologists (HK and IK). Patients were excluded if they had received high-dose steroids, intravenous immunoglobulin (IVIG), or plasma exchange within three months before screening or concurrent immunosuppressive therapy. We also excluded patients with a body mass index (BMI) of 40.

‘Early repleters’ were identified based on prior flow cytometry results obtained as part of clinical care. We defined a patient as an ‘Early Repleter’ (ER) if the percentage of CD19+ B cells in blood was >0.5% within 6 months after anti-CD20 infusion or >1.5% within 7 months after anti-CD20 infusion. Where possible, early repleters were matched to ‘Normal Repleter’ (NR) – patients who did not meet criteria for early repletion – based on age, sex, type of anti-CD20 therapy (ocrelizumab, OCR, or rituximab, RTX), duration of anti-CD20 therapy and BMI.

### Sample processing

Peripheral blood samples were collected from all participants during the bi-annual clinic visit that was scheduled before the anti-CD20 infusion. Sera were separated, and peripheral blood mononuclear cells (PBMCs) were isolated by density-gradient centrifugation. Isolated PBMCs were then aliquoted and cryo-stored in liquid nitrogen until further analysis.

### Flow cytometric analyses

PBMC samples were thawed, stained, and studied on the same day. Cells were stained with Live/Dead Fixable Blue Dead Cell Stain kit (ThermoFisher Scientific), and Fc receptors were blocked (Human TruStain FcX, BioLegend), as per manufacturer’s instructions. Surface staining was performed, and intracellular staining with FOXP3 Fix/Perm kit (ThermoFisher Scientific). Cells were washed fixed and data acquired on a 5-laser (355/405/488/561/637 nm) SONY ID7000 Spectral analyzer. 250,000 lymphocytes per sample were collected. Immune cell subsets assessed included naïve T cells (CD3+CD4+), regulatory T cells (CD3+CD25+CD127low), T peripheral helper cells (CD3+CD4+PD1+CXCR5-), classical monocytes (CD14+CD16-), non-classical monocytes (CD14-CD16+), and classical NK cells (CD56dim CD16+), as well as naïve B cells (CD19+CD27-IgD+), switched memory B cells (CD19+CD27+IgD-), unswitched memory B cells (CD19+CD27+IgD+), double negative (CD19+CD27-IgD-) B cells. Populations were quantitated based on established phenotyping criteria. Data acquisition was performed using the FACSDiva (Becton-Dickinson), with analysis using FlowJo (v10.10) software. The gating strategy is shown in Supplemental Figure 1. All surface markers, fluorophores, and antibody clones are listed in Supplemental Table 1. All data was reviewed by a board-certified hematopathologist (AAA).

### Serologic immunoassays testing

Serum B-cell activating factor (BAFF) (R&D Labs, PA), soluble CD40 ligand (sCD40L) (R&D Labs), and B-cell maturation antigen (BCMA) (R&D Labs) levels were measured by ELISA, according to the manufacturer’s instructions. In the OCR-treated patients with rapid repletion of B cell levels after infusion, serum OCR and anti-OCR levels were evaluated. In RTX-treated ER patients, anti-RTX antibody levels (Sanguin, NL) were tested. Serum immunoglobulin quantitations were performed by ELISA with comparison to standard curves (Jackson Immunoresearch).

### Statistical Analyses of Genetic SNP

Genetic variation was analyzed using the Infinium Immunoarray-24 v2 BeadChip (Illumina), a genotyping array that detects 247,814 single nucleotide polymorphisms (SNPs) related to immune system processes. To mitigate the high risk of false-positive findings inherent in assessing this large number of SNPs across our limited sample size, we implemented a multi-faceted approach. In our analyses, only SNPs present in at least two early repleter (ER) patients were considered for further analysis of representation. Subsequently, we used g:SNPense (g:Profiler)^13^ to map SNPs to gene names and assess these variants’ potential effects on gene function, as defined by Sequence Ontology. To further identify potentially relevant SNPs, we employed Fisher’s exact tests for association analysis, and we calculated odds ratio (OR) confidence intervals for effect size estimation. We also compared the mean allele frequencies between ER and NR groups. This multi-step approach, outlined in Supplemental Figure 2, aimed to identify biologically relevant SNPs while minimizing the risk of spurious associations.

Comparative analyses of demographic variables and clinical characteristics between early repleter (ER) and normal repleter (NR) patients were performed using chi-square tests for categorical variables and non-parametric t-tests (Mann-Whitney U testing) for continuous variables. Fisher’s exact test was applied to assess the association between SNPs and repletion status (ER vs. NR) in a 2 x 3 table of germline heterozygous and homozygous alleles. Odds ratios (ORs) were calculated for homozygous vs. germline and heterozygous vs. germline SNPs. SNPs of particular interest were those that had a differential overrepresentation in ER vs. NR, as defined by Fisher’s exact test p-value <0.05 and a lower confidence interval >1 for the odds ratio. All statistical analyses and visualizations were conducted using GraphPad Prism (Version 10.2.3) or R software (Version R 4.3.3).

### Pathway Analyses

To identify genes related to immune response and B cell differentiation that are significantly enriched in the ER group, pathway analysis was conducted using g:Profiler^10^. Hypergeometric calculations were used to determine the significance of gene set overlaps. A rank-ordered list of genes, which were identified based on the SNPs overrepresented in the ER group, was entered into the g:Profiler algorithm. In ER patients, top priority was assigned to genes associated with more than one SNP overrepresented, with a lower ranking based on the highest minor allele frequency (MAF). This approach enabled the systematic identification of gene SNP-associated pathways involved in immune system responses. To consider the relevance of a gene SNP for B cell development and differentiation, we used a reference list of 8803 genes compiled from reports that defined genes differentially expressed during critical stages of B cell development^14-17^.

## Results

### Patient characteristics and disease course in normal repleters (NR) and early repleters (ER)

We enrolled 20 AA MS patients who met our criteria for ER. All were diagnosed with MS, 80% were female, the mean age was 38.7 years [SD=12.9], mean duration of anti-CD20 therapy was 4.9 years [range: 2 - 8 years]. We enrolled 23 AA patients who were NR, of whom 21 had MS and 2 had NMO, 87% were female, the mean age was 42.3 ± 9.9 years, and the mean duration on anti-CD20 therapy was 4.9 years [range: 1.3 - 14 years]. As shown in Table 1, there were no differences in the demographic and clinical characteristics of the two groups.

**Table 1.**
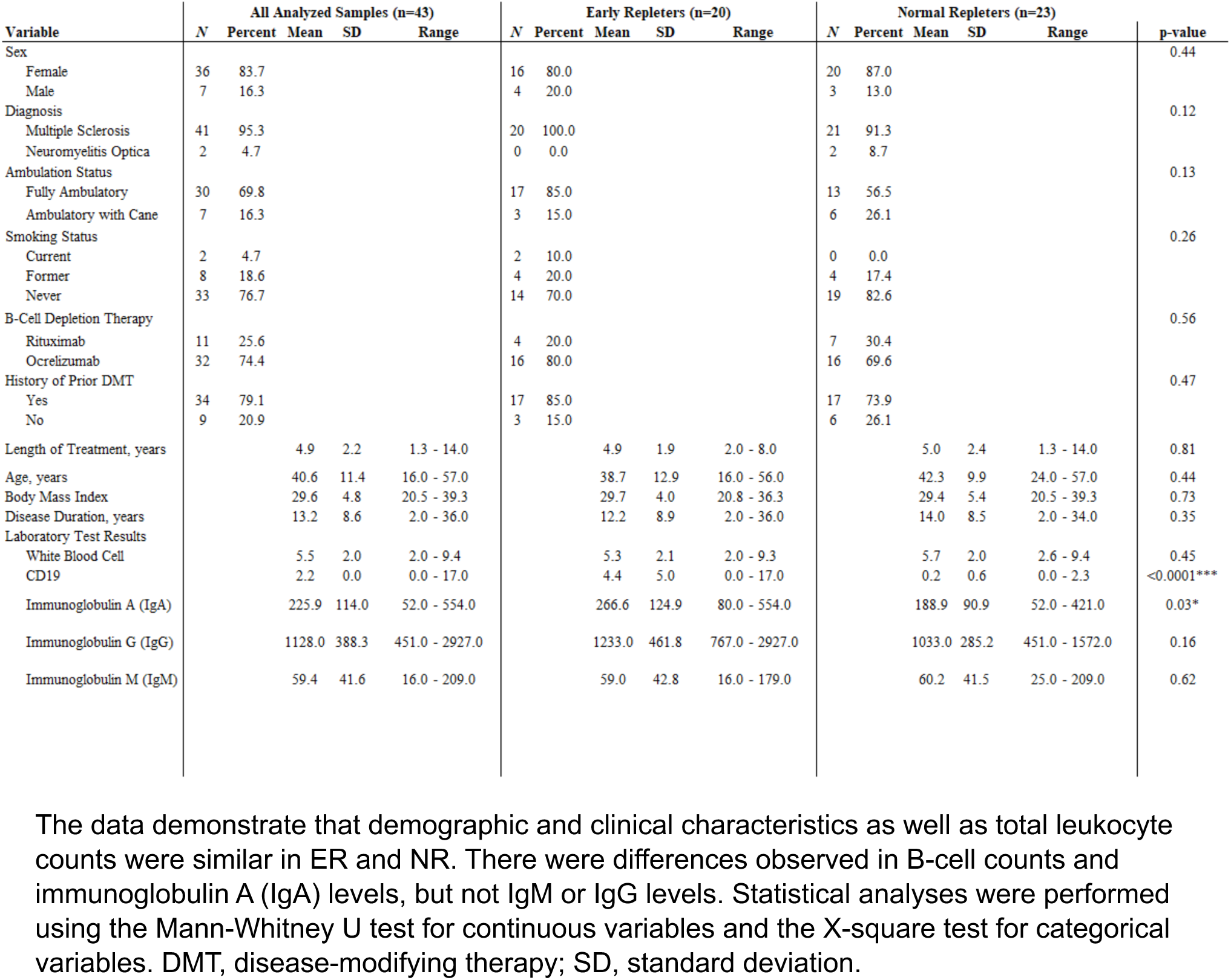
Comparative analysis of demographics variables, leukocyte and B-cell (CD19+) counts, and immunoglobulin levels in early repleters (ER) and normal repleters (NR).

| Variable | All Analyzed Samples (n=43) |  |  |  |  | Early Repleters (n=20) |  |  |  |  | Normal Repleters (n=23) |  |  |  |  | p-value |
| --- | --- | --- | --- | --- | --- | --- | --- | --- | --- | --- | --- | --- | --- | --- | --- | --- |
|  | N | Percent | Mean | SD | Range | N | Percent | Mean | SD | Range | N | Percent | Mean | SD | Range |  |
| Sex |  |  |  |  |  |  |  |  |  |  |  |  |  |  |  | 0.44 |
| Female | 36 | 83.7 |  |  |  | 16 | 80.0 |  |  |  | 20 | 87.0 |  |  |  |  |
| Male | 7 | 16.3 |  |  |  | 4 | 20.0 |  |  |  | 3 | 13.0 |  |  |  |  |
| Diagnosis |  |  |  |  |  |  |  |  |  |  |  |  |  |  |  | 0.12 |
| Multiple Sclerosis | 41 | 95.3 |  |  |  | 20 | 100.0 |  |  |  | 21 | 91.3 |  |  |  |  |
| Neuromyelitis Optica | 2 | 4.7 |  |  |  | 0 | 0.0 |  |  |  | 2 | 8.7 |  |  |  |  |
| Ambulation Status |  |  |  |  |  |  |  |  |  |  |  |  |  |  |  | 0.13 |
| Fully Ambulatory | 30 | 69.8 |  |  |  | 17 | 85.0 |  |  |  | 13 | 56.5 |  |  |  |  |
| Ambulatory with Cane | 7 | 16.3 |  |  |  | 3 | 15.0 |  |  |  | 6 | 26.1 |  |  |  |  |
| Smoking Status |  |  |  |  |  |  |  |  |  |  |  |  |  |  |  | 0.26 |
| Current | 2 | 4.7 |  |  |  | 2 | 10.0 |  |  |  | 0 | 0.0 |  |  |  |  |
| Former | 8 | 18.6 |  |  |  | 4 | 20.0 |  |  |  | 4 | 17.4 |  |  |  |  |
| Never | 33 | 76.7 |  |  |  | 14 | 70.0 |  |  |  | 19 | 82.6 |  |  |  |  |
| B-Cell Depletion Therapy |  |  |  |  |  |  |  |  |  |  |  |  |  |  |  | 0.56 |
| Rituximab | 11 | 25.6 |  |  |  | 4 | 20.0 |  |  |  | 7 | 30.4 |  |  |  |  |
| Ocrelizumab | 32 | 74.4 |  |  |  | 16 | 80.0 |  |  |  | 16 | 69.6 |  |  |  |  |
| History of Prior DMT |  |  |  |  |  |  |  |  |  |  |  |  |  |  |  | 0.47 |
| Yes | 34 | 79.1 |  |  |  | 17 | 85.0 |  |  |  | 17 | 73.9 |  |  |  |  |
| No | 9 | 20.9 |  |  |  | 3 | 15.0 |  |  |  | 6 | 26.1 |  |  |  |  |
| Length of Treatment, years |  |  | 4.9 | 2.2 | 1.3 - 14.0 |  |  | 4.9 | 1.9 | 2.0 - 8.0 |  |  | 5.0 | 2.4 | 1.3 - 14.0 | 0.81 |
| Age, years |  |  | 40.6 | 11.4 | 16.0 - 57.0 |  |  | 38.7 | 12.9 | 16.0 - 56.0 |  |  | 42.3 | 9.9 | 24.0 - 57.0 | 0.44 |
| Body Mass Index |  |  | 29.6 | 4.8 | 20.5 - 39.3 |  |  | 29.7 | 4.0 | 20.8 - 36.3 |  |  | 29.4 | 5.4 | 20.5 - 39.3 | 0.73 |
| Disease Duration, years |  |  | 13.2 | 8.6 | 2.0 - 36.0 |  |  | 12.2 | 8.9 | 2.0 - 36.0 |  |  | 14.0 | 8.5 | 2.0 - 34.0 | 0.35 |
| Laboratory Test Results |  |  |  |  |  |  |  |  |  |  |  |  |  |  |  |  |
| White Blood Cell |  |  | 5.5 | 2.0 | 2.0 - 9.4 |  |  | 5.3 | 2.1 | 2.0 - 9.3 |  |  | 5.7 | 2.0 | 2.6 - 9.4 | 0.45 |
| CD19 |  |  | 2.2 | 0.0 | 0.0 - 17.0 |  |  | 4.4 | 5.0 | 0.0 - 17.0 |  |  | 0.2 | 0.6 | 0.0 - 2.3 | <0.0001*** |
| Immunoglobulin A (IgA) |  |  | 225.9 | 114.0 | 52.0 - 554.0 |  |  | 266.6 | 124.9 | 80.0 - 554.0 |  |  | 188.9 | 90.9 | 52.0 - 421.0 | 0.03* |
| Immunoglobulin G (IgG) |  |  | 1128.0 | 388.3 | 451.0 - 2927.0 |  |  | 1233.0 | 461.8 | 767.0 - 2927.0 |  |  | 1033.0 | 285.2 | 451.0 - 1572.0 | 0.16 |
| Immunoglobulin M (IgM) |  |  | 59.4 | 41.6 | 16.0 - 209.0 |  |  | 59.0 | 42.8 | 16.0 - 179.0 |  |  | 60.2 | 41.5 | 25.0 - 209.0 | 0.62 |

The data demonstrate that demographic and clinical characteristics as well as total leukocyte counts were similar in ER and NR. There were differences observed in B-cell counts and immunoglobulin A (IgA) levels, but not IgM or IgG levels. Statistical analyses were performed using the Mann-Whitney U test for continuous variables and the X-square test for categorical variables. DMT, disease-modifying therapy; SD, standard deviation.

Neurologist-diagnosed relapses while on anti-CD20 therapy occurred in 4/20 ER patients (20%); the mean annual relapse rate (ARR) of ER was 4.1%. Relapses while on anti-CD20 therapy were recorded for 5 NR/23 patients (22%), with one patient having 2 relapses; the mean annual relapse rate in NR was 5.3%, which is similar to ARR of ER patients (p=0.849). In ER patients, a total of 88 MRIs were obtained while on anti-CD20 therapy, and in NR patients - 91 MRIs. New MRI lesions were observed in 4/20 ER patients (20%; mean annual lesion formation rate of 4.1%) and in 3/23 NR patients (13%; mean annual lesion formation rate of 2.7%). There was no difference between new lesion rates in the two groups (p=0.796). Clinical and MRI activity for each of the participants throughout anti-CD20 therapy is shown in Supplemental Figure 3.

ER and NR patients had similar time from the anti-CD20 infusion to blood sample collection (p=0.3) (Table 1). As expected, ER patients had significantly higher levels of CD19+ B cells compared to the NR (p <0.0001, Table 1). All ER patients had detectable levels of B-cells within 6 months post-infusion, while none of the NR patients had detectable circulating B cells within 6 months of their last anti-CD20 infusion (Figure 1). Serum IgA levels were significantly higher in the ER group compared to the NR group (p = 0.043), while serum IgG and IgM levels were not different (p = 0.35, p = 0.61, respectively) (Table 1).

**Figure 1.**
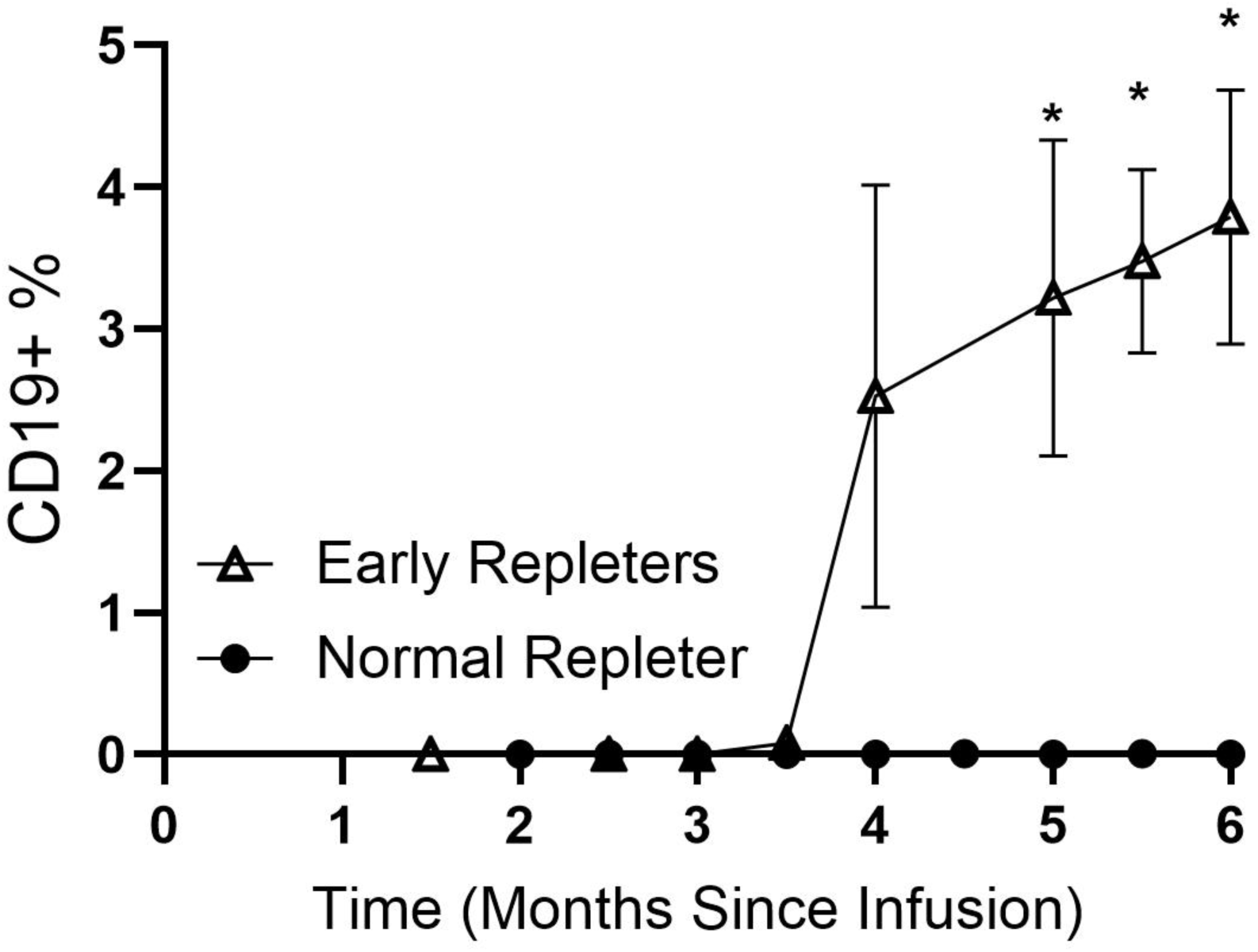
B cell repopulation post-CD20 depletion in early repleters (ER) and normal repleters (NR). Data for 23 NR and 20 ER MS patients are shown, with a total of 133 longitudinal samples collected over 1.5 to 6 months post anti-CD20 infusion. Values for CD19+ (B lineage) cells are percentages of total circulating lymphocytes. Data are presented as mean ± standard error of the mean. * indicates the specific time points attaining p<0.01 (Mann-Whitney U test).

### Anti-CD20 concentrations and anti-drug levels in ER and NR patients

To ascertain whether differences in rates of B-cell repletion in the two groups may be due to differences in drug bioavailability, we assessed OCR concentration levels in a subset of 17 OCR-treated patients (6 NR and 10 ER). There were no significant differences in the number of days between the blood collection and the last OCR infusion in the NR- and ER-tested patients (NS, p=0.62) and no differences in OCR concentrations in NR- and ER-tested patients (Supplemental Figure 4A&B).

We also assessed for the presence of anti-drug antibodies (ADA) (i.e., anti-OCR antibodies) in the same 17 patients and found detectable ADA levels in two OCR-treated ER patients. The two patients with anti-OCR ADA had no detectable OCR drug levels (<0.0025 ug/mL). B-cell repletion histories for the two patients with anti-OCR ADA are shown in Table 2.

**Table 2.** B cell repletion in patients with anti-ocrelizumab antibodies.

| Anti-OCR Antibody (AU/mL) | OCR Level (ug/mL) | % B Cell Repletion | Total IgG level (mg/dL) | Time Since Infusion (Months) | Additional Information |
| --- | --- | --- | --- | --- | --- |
| 576 | <0.0025 | 17% | 3279 | 3-4 | - Previous treatments: Fingolimod<br>- Co-morbidities: SLE and Sjogren's syndrome |
|  |  | 13% | 2952 | 4-5 |  |
|  |  | 14% | 2917 | 6-7 |  |
| 101 | <0.0025 | 4% | 1728 | 4-5 | - Previous treatments: Interferon beta-1a<br>- Co-morbidities: Hyperlipidemia |
|  |  | 5.5% | 1490 | 7-8 |  |
|  |  | 5.5% | 1615 | 8-9 |  |

Ten ER and six NR were among the participants who were tested for anti-ocrelizumab anti-drug antibodies (ADA), no NR participant has ADA, while two ER patients with ADA were identified. These two patients with detectable levels of anti-ocrelizumab antibodies (>87.5 AU/mL) are highlighted in the table. The levels of these antibodies and the available post-infusion level of circulating CD19+ cells are provided for each patient. OCR, ocrelizumab.

**Table 3.**
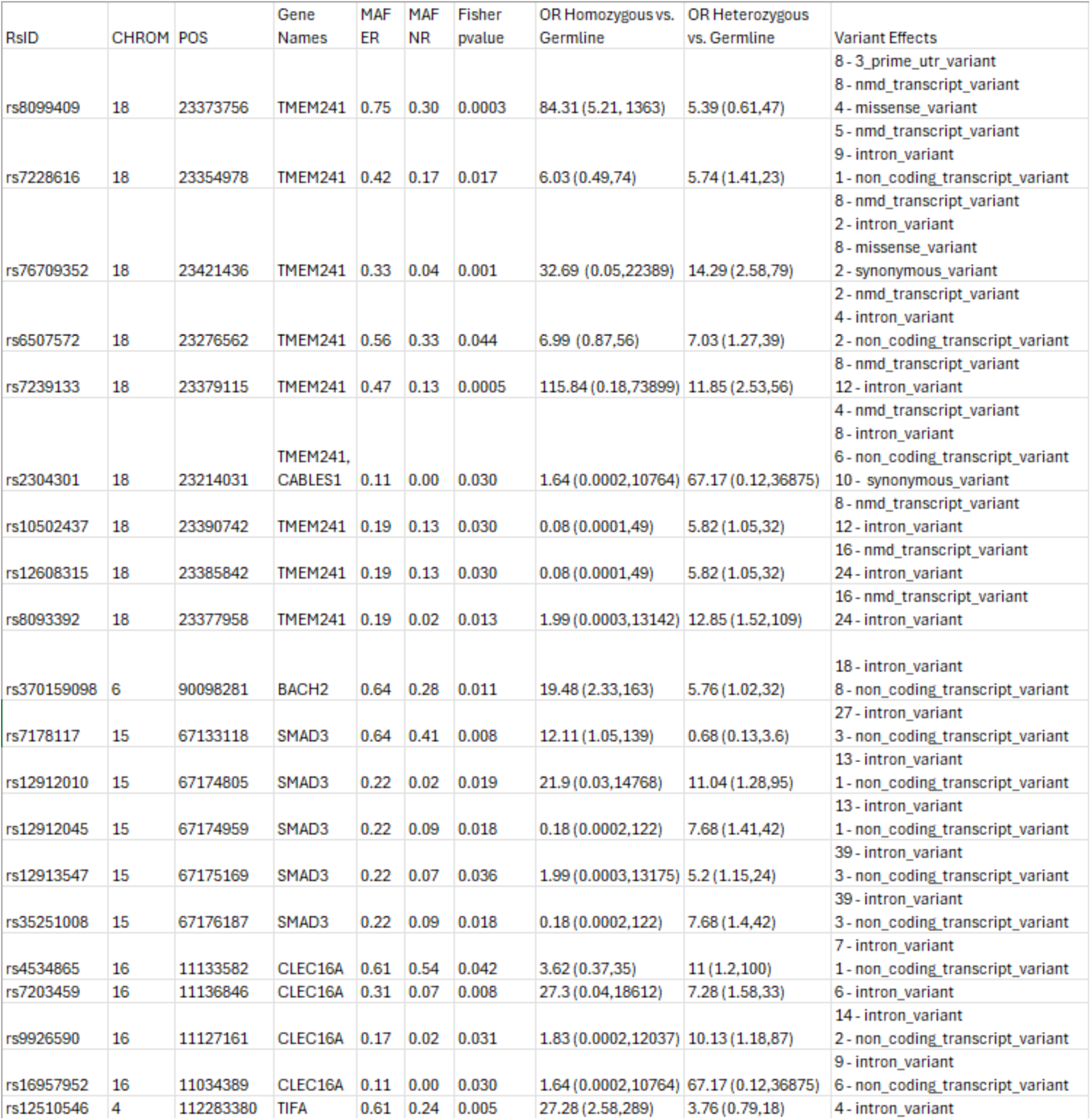
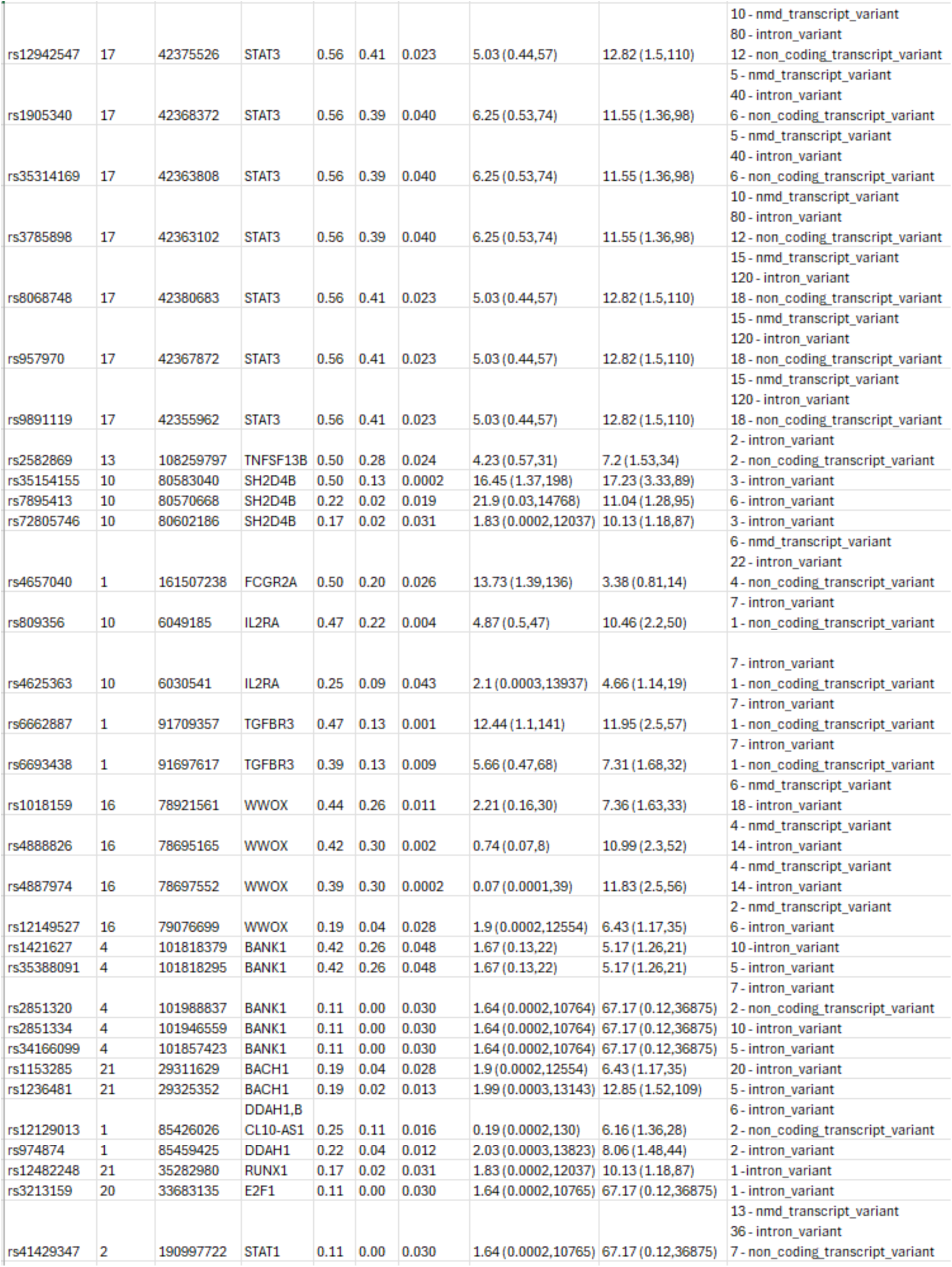
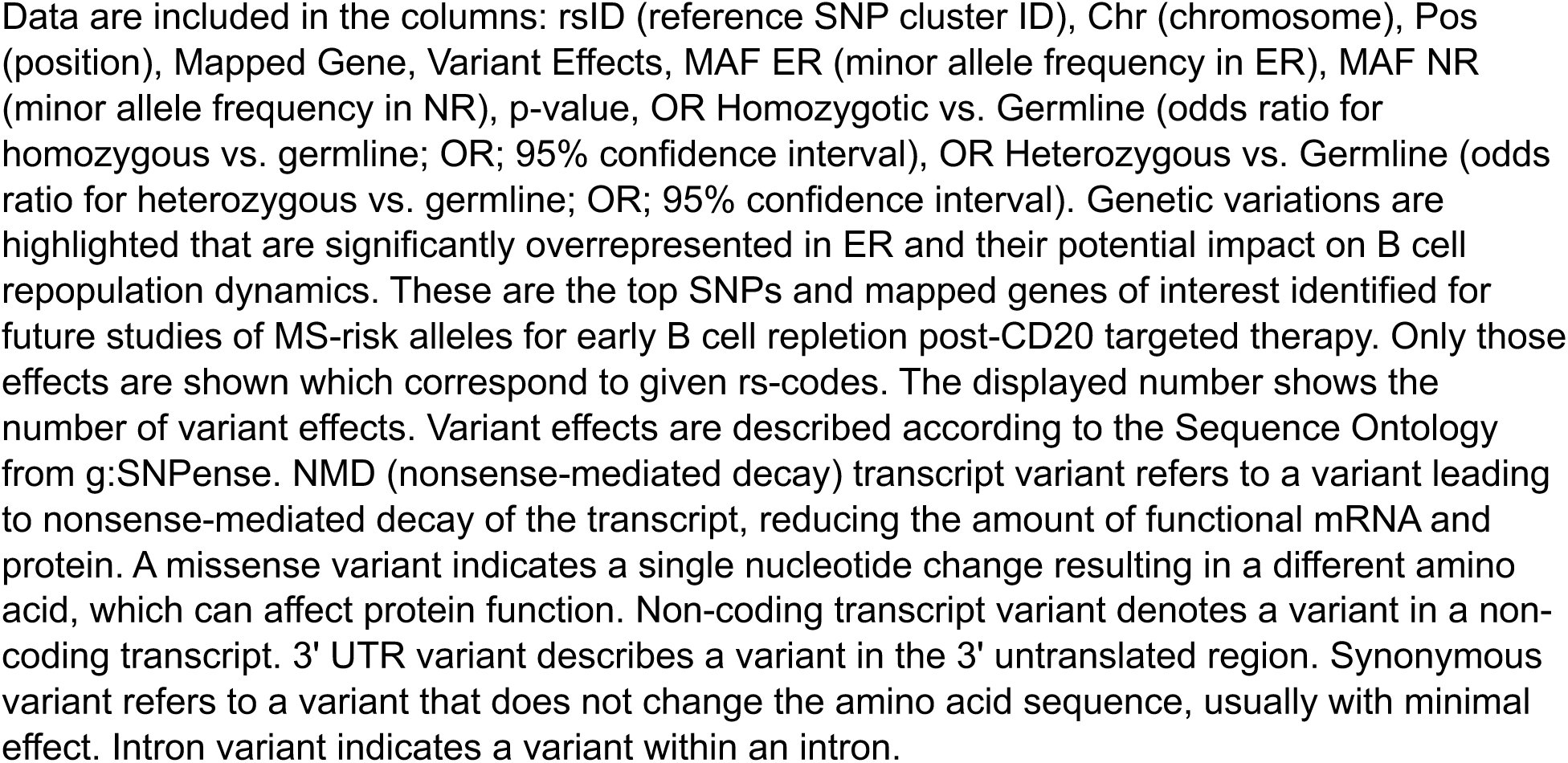
SNPs are overrepresented in Early Repleters (ERs) compared to Normal Repleters (NRs) and the association with MS risk genes.

Among the OCR-treated patients without ADA, there was no correlation between blood CD19+ B cell level and OCR (Supplemental Figure 4C). Three out of 4 of the RTX-treated ER patients were available for anti-RTX antibody testing, and none had ADA.

Levels of B-cell survival factors but not activation factors differ between NR and ER patients Soluble factors can have dramatic systemic effects on B cell survival, differentiation, and activation, and we, therefore, examined serum levels of the TNF family member co-stimulation factor, CD40 ligand (CD40L), as well as the soluble form of the B cell maturation antigen (BCMA) that is shed by plasma cells. We found that these levels were similar in the ER and NR groups (Figure 2).

**Figure 2.**
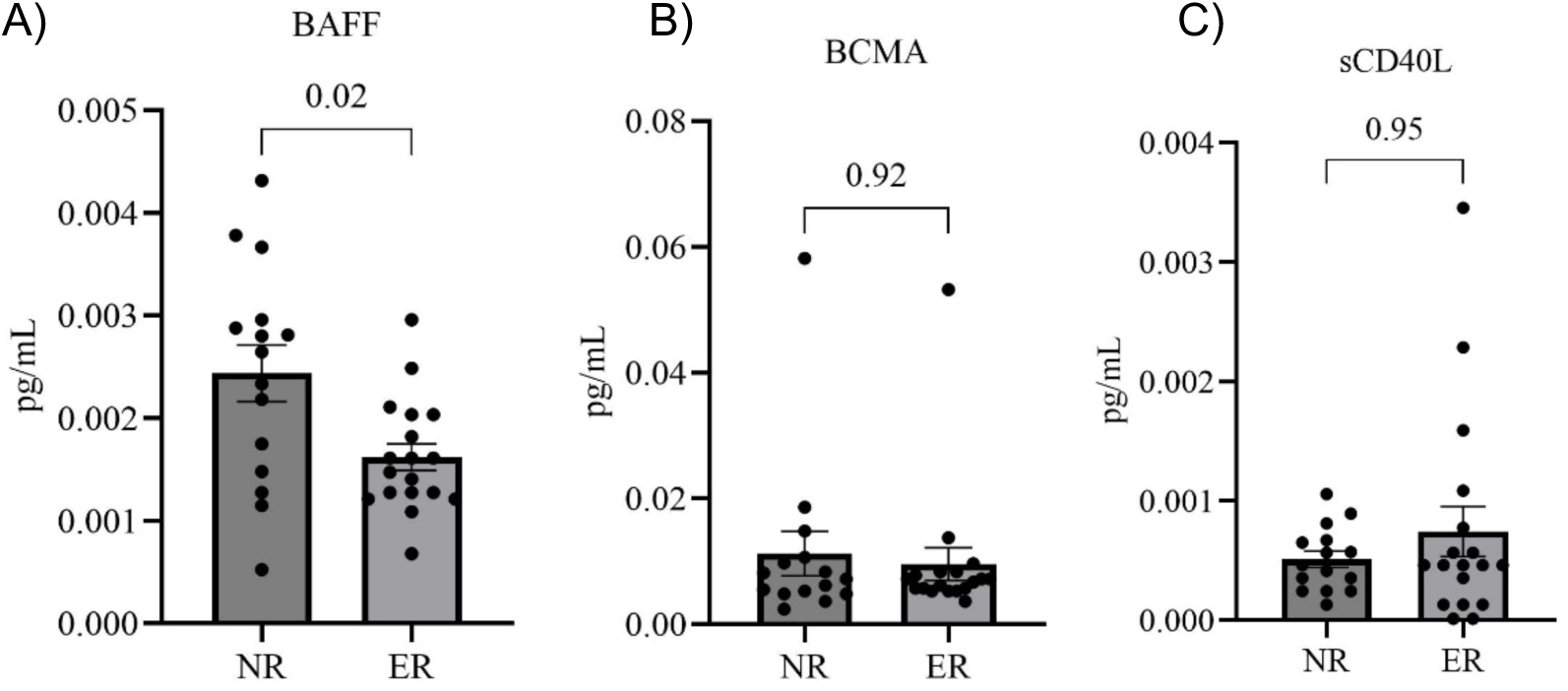
Serum levels of BAFF, sCD40L, and BCMA in normal repleters (NR, n=15) vs. early repleters (ER, n=18). A) Serum levels of B-cell activating factor (BAFF) are significantly higher in the NR group (p=0.02). B) Soluble CD40 ligand (sCD40L) levels were not different, and C) B-cell maturation antigen (BCMA) were also not different between ER and NR groups. All serum samples were drawn between 3 and 6 months post-ocrelizumab treatment. Quantification was performed using commercial ELISA. Statistical comparisons were made using the Mann-Whitney U test. Data represent mean ± standard error of the mean.

Based in part on a report of an MS cohort with overexpression of specific BAFF-related polymorphism that could lead to B-cell dysregulation^18^, we next examined levels of B cell activation factor (BAFF), also termed B lymphocyte stimulator, that affects the survival of cells of the B lineage. We found serum BAFF levels were significantly higher in NR than in ER patients (p = 0.02) (Figure 2). This pattern was expected, as reciprocal systemic BAFF elevations are known to be concurrent with the overall depletion of B-lineage cells in the body^19^. The lower BAFF levels in ER patients likely reflect the greater consumption of BAFF because of more rapid repopulation and differentiation of the peripheral B-cell pool and, therefore, serve as independent confirmation of early repleter status.

### Representation of B cell subsets in ER and NR groups

Complete blood cell counts were assessed at a mean of 4.9 (± 1.9) months after the last anti-CD20 infusion. All samples contained more than 1 x 10^9 total lymphocytes/mL. There were no significant differences in the peripheral blood percentages of naive CD4+ T cells, naïve CD8+ T cells, regulatory T cells (CD4+CD25+), or T peripheral helper cells (TPH) (CD4+PD1+CXCR5-) between the ER and NR groups (Supplemental Figure 5).

We also examined the representation of B lymphocyte subsets, defined as CD19-bearing, in peripheral blood samples collected after anti-CD20 infusions. Notably, B cell repletion was higher in the bloodstream of the ER group (Figure 1), but the representation of the B cell subsets was comparable, with no significant differences in the percentages of naive mature (CD27-sIgD+), switched memory (CD27+IgD-), unswitched memory (CD27+IgD+), or Double Negative (CD27-sIgD-) B cells in the ER compared to the NR group, relative to the total number of B cells, which suggested these two groups displayed similar repopulation dynamics. In both ER and NR, transitional and immature/naïve B cells, which are the subsets that newly arise to enter the circulation, were the dominant subsets (>95% in both NRs and ERs) (Supplemental Figure 5).

### Genetic variation and SNP analysis

We next investigated for differences in the inheritance genetic variants with possible involvement in early B cell repopulation. For the genetic analyses, the two patients with raised anti-OCR antibodies were omitted, as immune-mediated B cell clearance mechanisms were sufficient to account for earlier B-cell repletion in these two subjects. We explored the genetic factors that potentially contribute to early B cell repletion in AA MS, using the Immunoarray Beadchip (Illumina) and performing case-control association analysis. Out of the 247,814 SNPs tested, 189,777 SNPs were found in at least two of the ER patients in our cohort. Of these, there was a significant overrepresentation of 6,471 SNPs in the ER. Furthermore, of these, 3,724 had a higher mean allele frequency in ER (Fisher’s exact test, p < 0.05). We further determined the representation of homozygous and heterozygous SNPs in ER versus NR. This analysis identified 1,887 SNPs of interest (1,520 heterozygous vs. germline; 281 homozygous vs. germline; 84 both homozygous and heterozygous vs. germline) with the lower limit of the confidence interval for the odds ratios ≥1. Additionally, among the significantly overrepresented SNPs (Fisher’s exact test, p < 0.005) with a positive mean allele frequency in ERs compared to NRs, we identified 317 SNPs of interest that had a mean allele frequency of zero in NRs but an allele frequency of at least 2 in ERs (Supplemental Table 2).

The distribution of SNPs across the genome is shown in the Manhattan plot (Figure 3). Notably, four of the 2,202 SNPs overrepresented in ER, with a p-value <0.001, were associated with genes TGFBR3, SH2D4B, TMEM241, and WWOX, which are known to have roles in immune system function, including B cell development. A volcano plot illustrating the odds ratios for homozygous versus germline and heterozygous versus germline SNPs in ER versus NR is shown (Figure 4). Of SNPs of interest, the mean allele frequency was significantly higher in ERs than in NRs (Figure 5). Notably, these SNPs also had a higher mean allele frequency (MAF) in ERs compared to reference control subjects of African descent described by Illumina (Supplemental Figure 6A&B).

**Figure 3.**
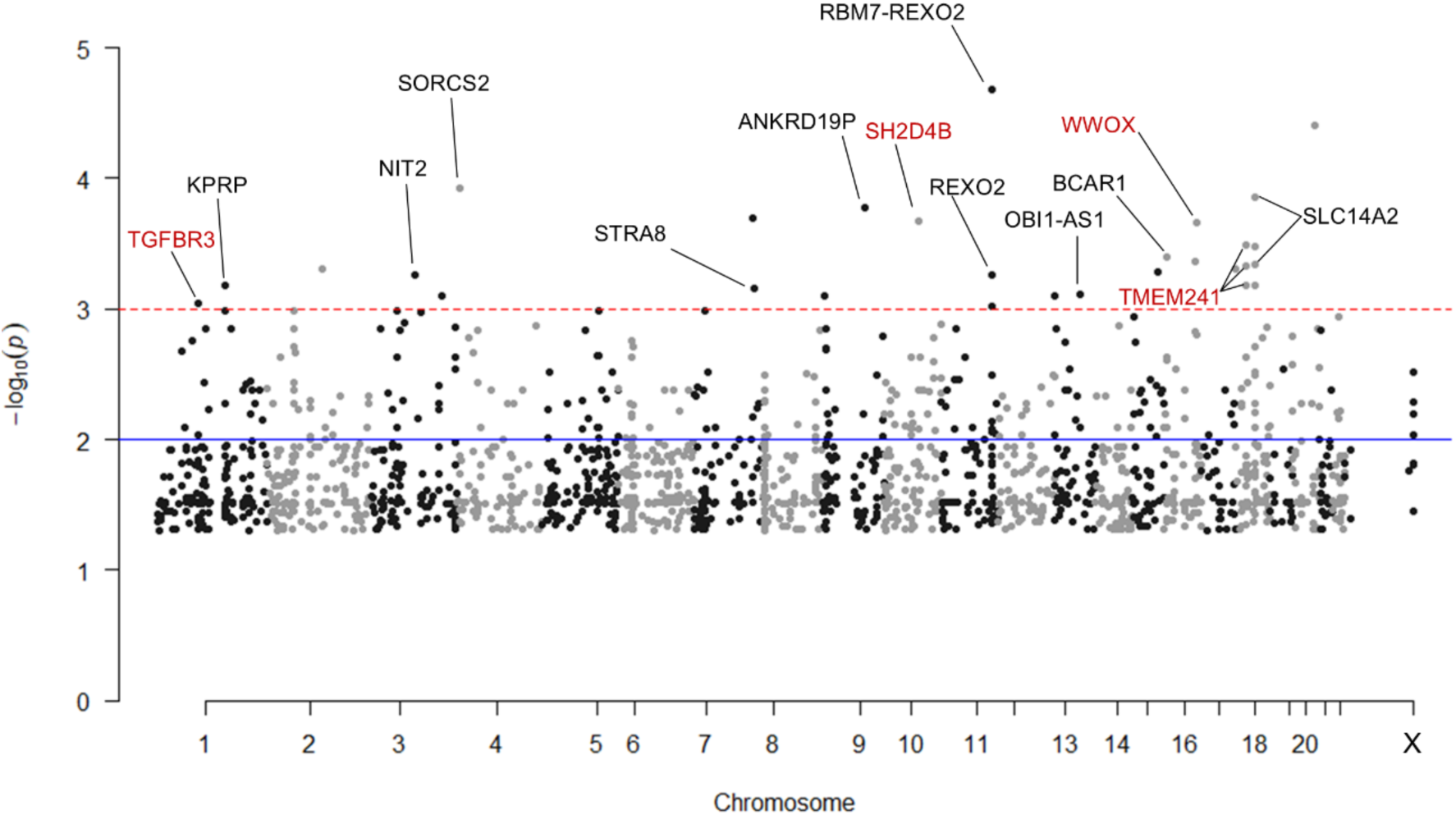
Manhattan plot of 2,202 SNPs significantly overrepresented in early repleters (ER) vs. normal repleters (NR). The plot displays SNPs identified as significantly overrepresented among ER participants by Fisher’s exact test (p-value < 0.05) and meeting at least one of three additional significance criteria: 1) Lower limit confidence interval > 1 for the odds ratio of homozygous to germline SNPs, 2) Lower limit confidence interval > 1 for the odds ratio of heterozygous to germline SNPs, 3) Mean allele frequency of for NRs with at least 2 SNP alleles identified in ERs. SNPs with a negative log10(p-value) > 3 that are mapped to genes by g:SNPense are annotated. Gene names in red indicate SNPs of interest related to immune system processes, particularly B cell activation and development.

**Figure 4.**
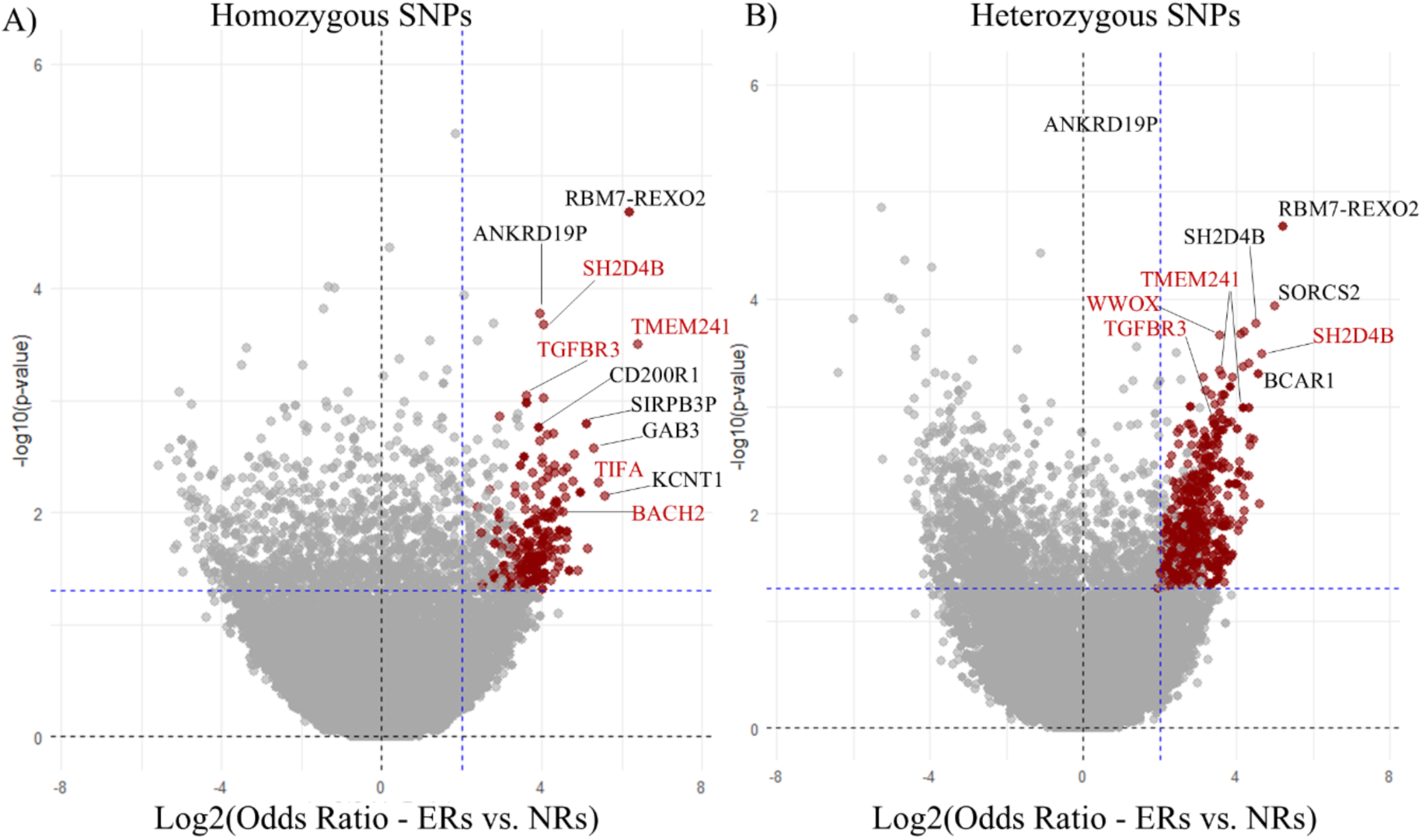
Odds ratios of SNPs in early repleters (ER) vs. normal repleters (NR). This figure presents the odds ratios of homozygous (A) and heterozygous (B) SNPs when comparing early repleters (ER) to normal repleters (NR). SNPs highlighted in red indicate those with a Fisher’s exact test p< 0.05, and an odds ratio lower limit >1 (365 for homozygous and 1,604 for heterozygous). The most overrepresented SNPs of interest are annotated with the names of their corresponding mapped genes (g:SNPense).

**Figure 5.**
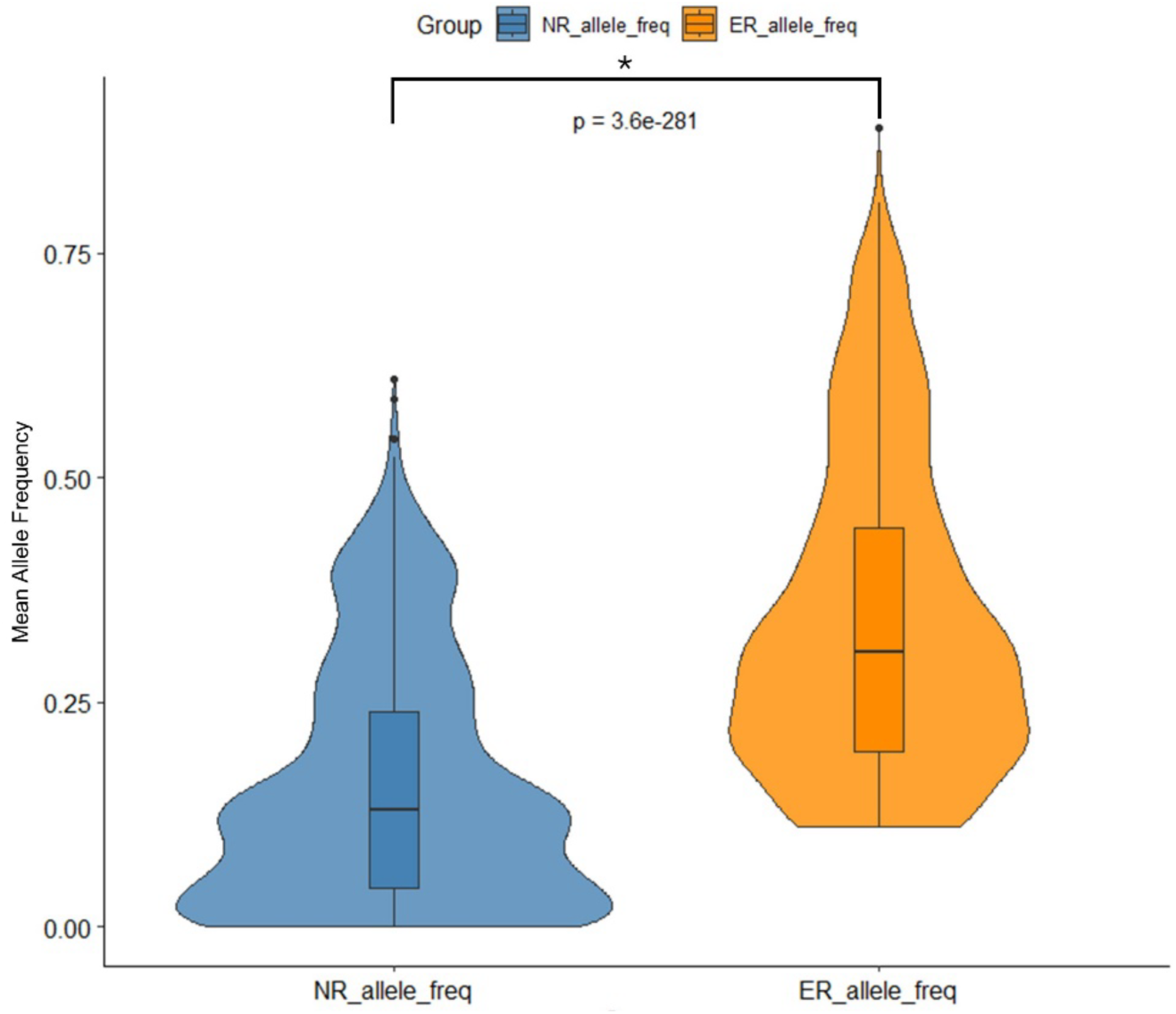
Distribution of allele frequencies in early repleter (ER) and normal repleter (NR) groups. The figure depicts the distribution of allele frequencies for 2,202 SNPs that were significantly overrepresented in the ER group. Violin plots show the density of allele frequencies, while the box plots indicate the median, quartiles, and potential outliers. Box and whisker plots represent the interquartile range (IQR), with the whiskers extending to 1.5 times the IQR from the first and third quartiles. Outliers are represented as individual points beyond the whiskers.

### Pathway analysis for SNPs with known gene variant effects

To investigate potential functional relationships between SNPs that are more common in ER, we completed a pathway analysis of SNPs with known variant effects on mapped genes (Figure 6). Using g:Profiler, we assessed pathway enrichment of genes affected by SNPs overrepresented in ER patients. SNPs overrepresented in ERs mapped to genes enriched in pathways related to cell surface receptor signaling (adjusted p =5.94E-05, normalized enrichment score (NES) = 13.1), intracellular signal transduction (adjusted p =0.02, NES =12.0), immune response (adjusted p =0.002, NES =10.7), leukocyte activation (adjusted p-value=3.06E-6, NES=5.08), inflammatory response (adjusted p=0.04, NES=4.5), lymphocyte activation (adjusted p=1.7E-5, NES=4.2), and cytokine production (adjusted p=1.46E-07, NES=4.2) (Figure 6). We also specifically assessed whether there was an overrepresentation of SNPs in ERs from genes known to play roles in B cell differentiation^14-17^. Our analysis identified an overrepresentation of 228 genes with known B cell differentiation-related function (adjusted p-value=1.9E-262, NES=12.5). Notably, these genes included BACH2, IRF1, and other key transcription factors critical for B cell development, as well as TNFSF13B, the gene encoding BAFF itself. Supplemental Table 3 lists the genes that map to known functional pathways related to SNPs overrepresented in ER.

**Figure 6.**
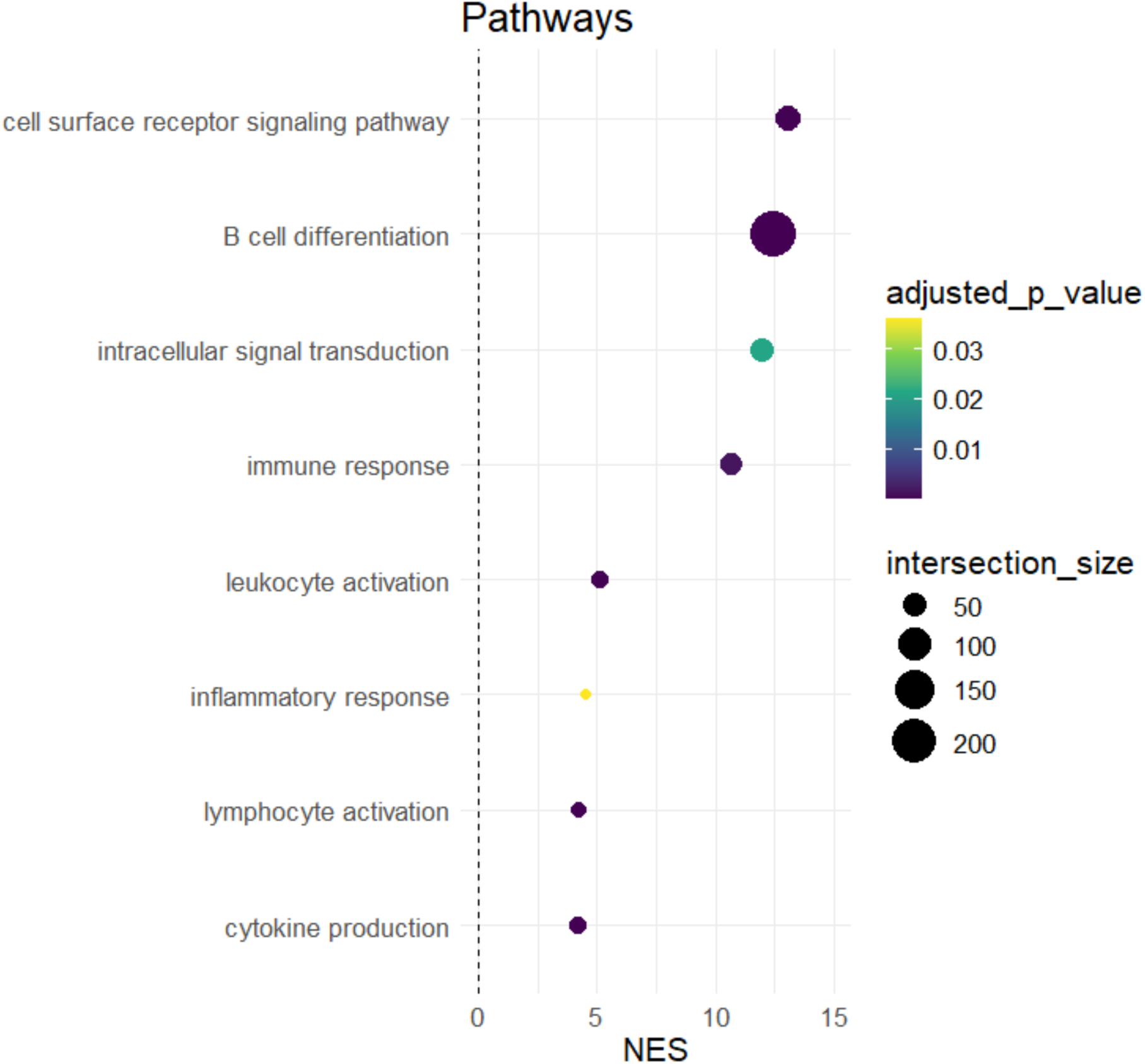
Pathway analysis of genes mapped to by SNPs overrepresented in early repleters (ER). The figure shows the results of a hypergeometric analysis indicating enrichment for pathways related to immune system processes and B cell differentiation. Colors represent adjusted p-values, and the size of the dots represents the number of genes included in the noted pathway.

### SNPs associated with the most rapid B-cell repletion

Within our group of 18 ER without ADA, we identified a subgroup of 7 patients, whom we termed "super-repleters,” as each displayed circulating B cell levels of >2% at 4 months after anti-CD20 therapy. In the seven super-repleters, 45 SNPs were significantly overrepresented compared to other ER patients (Fisher’s exact test, p < 0.05), and the higher minor allele frequency (MAF) in these super-repleters was also significantly overrepresented in ER compared to NR, including genes enriched for immune receptor activity; IL17RE, IL2RB, IL17RD, IL1R1, IL23R, and IL1RL2 (Supplemental Table 4), all related to inflammatory pathway.

We wondered whether differences in genetic inheritance could contribute to variations in levels of soluble CD40L (sCD40L) that arise via cleavage from the lymphocyte membrane. We therefore dichotomized the ER patients based on serum sCD40L levels: those with high sCD40L (n = 4) and those with low sCD40L (n = 14), with high levels defined as greater than the mean + SEM, as shown in Figure 2. For the ERs with high levels of serum sCD40L, we identified 27 SNPs that were significantly overrepresented compared to the representation in ER patients with low sCD40L (Fisher’s exact test p < 0.05). In the high sCD40L ER patients, the genes with higher MAF were also overrepresented in NR vs. ER comparisons, and this set included genes involved in interleukin-12-related signaling: PLCB1, IL12RB2, and STAT4 (Supplemental Table 5). Compared to the low sCD40L ER patients, the high sCD40L ER patients also had an overrepresentation of an SNP associated with an intron variant in the transcription factor, Bach1, which regulates B cell development and differentiation^20^.

### Alleles are over-represented in ER patients also associated with MS disease risk

In our cohort, we also examined genes previously identified as non-MHC risk alleles for MS in AA patients^21^. Among the assessed rsIDs, significant associations were observed for seven SNPs, previously described by Isobe et al. ^21^, that were overrepresented in both ER patients compared to NR in our cohort. These genes include WWOX, DDAH1, STAT3, CLEC16A, IL2RA, BACH2, and PHGDH. Notably, rs12149527, which has been associated with altered transcript half-life of the gene WWOX, was previously identified by Isobe et al.^21^ as an MS genetic susceptibility factor in AA individuals. In our study, WWOX was also associated with ER. Such effects on transcript levels can lead to altered expression, potentially impacting the gene’s function as a susceptibility gene for MS. Hence, mechanisms predisposing AA individuals to develop MS may overlap with mechanisms that are responsible for early B cell repopulation, suggesting that pathologic B cell autoimmunity may also be intertwined with the drivers of faster B cell repopulation after anti-CD20 infusion.

## Discussion

Our investigations confirmed that amongst AA patients with neurologist-diagnosed MS, there is a distinct subpopulation, here termed ER, exhibiting a time-shortened B cell depletion response to standard semi-annual treatment dosing of anti-CD20 monoclonal antibodies. In both ER and NR, the repletion pattern was dominated by early B cell subsets (transitional and immature/naïve B cells), likely emerging after being newly generated in the bone marrow.

We assessed anti-CD20 drug concentrations and the presence of anti-drug antibodies. Although therapeutic anti-CD20 antibodies, including OCR, were engineered to be more human-like to subvert the development of anti-drug antibody responses, these still possess molecular features that are foreign to the human immune system. Yet, amongst the 43 AA MS patients studied, we identified early repleters, of which two were found to have high titers of anti-OCR antibodies (4.6%), a higher prevalence than the 0.4% reported in a clinical trial of diverse ethnicities and races^6^, though our population was in part selected based on their early B cell repopulation in prior treatment cycles. Predictably, the anti-OCR antibodies in our patients appeared to functionally neutralize and/or enhance OCR clearance, which consistently correlated with early post-infusion B cell repletion. Whether patients of African descent are more likely to develop ADA is a question that warrants further study.

Multiple lines of evidence suggest that inherent differences in B cell repopulation kinetics in individuals of African descent could influence their disease course in MS and other autoimmune diseases. AA patients with MS are reported to have a higher representation of B cells and plasmablasts in the cerebrospinal fluid^22^, and more commonly exhibit oligoclonal bands than patients of other race/ethnic groups^23,24^. Indeed, AA patients with systemic lupus erythematosus display higher serum BAFF levels and a greater number of circulating activated B cells^25-27^. Furthermore, even among healthy individuals, AA patients tend to demonstrate heightened neutralizing antibody responses to influenza vaccination^28,29^.

In our patients, we observed an appropriate reciprocal depression of BAFF levels concurrent with peripheral B cell replenishment. In MS patients, the differences in BAFF levels in ER and NR thus appear to be a consequence rather than the cause of early repletion. While lower BAFF levels in sera may, in part, be secondary to increased B cell repletion in ER patients, this does not rule out the role of SNPs on BAFF function and their potential pathogenic roles of B-cells in autoimmunity. TNFSF13B, which encodes the BAFF cytokine, plays a central role in peripheral B-cell survival and maturation. Notably, an intron variant and non-coding transcript variant SNP (rs2582869) impacting TNFSF13B transcript half-life (encoding a BAFF receptor) were overrepresented in ER patients, and this same SNP is also associated with the B cell cancer, non-Hodgkin’s lymphoma (NHL)^30^.

Cis-based cell signaling can have important survival effects on B cells, even when serum BAFF levels are unremarkable. Although the exact impact of rs2582869 on BAFF expression is uncertain, the SNP rs9514827 in the promoter of BAFF (-871C->T) has been associated with increased BAFF transcription ^33^. Notably, rs2582869 and rs9514827 are in the same genomic region, suggesting that genetic variations in this area may both have significant effects on BAFF function and resultant B cell activity. We also found overrepresentation of other SNPs relevant to immune pathways, including those with known effects on the following genes: IL4, STAT3, and NOD2 (inflammatory response), IL2RA, CD226, CCR6 (leukocyte activation), EBF1, BANK1, and BACH2 (B cell differentiation). Further investigation of genetic features identified in our study is therefore warranted in a larger cohort of AA patients to elucidate the mechanisms driving early B cell reconstitution.

We also found that the SNP rs465704, which maps to the FCGR2A gene, is significantly overrepresented in ER MS patients. This SNP is classified as both an NMD (nonsense-mediated decay) transcript variant and an intron variant (Supplemental Table 2), which may affect the half-life of the transcript. FCGR2A polymorphisms have been shown to affect the pharmacogenetics of rituximab by altering the cytotoxic function of macrophages and natural killer cells, thereby impacting the efficacy of monoclonal antibody therapies. Antibody-dependent cellular cytotoxicity (ADCC) plays a crucial role in the clinical effectiveness of rituximab in a range of disease states^31^. Research in other autoimmune diseases, including Neuromyelitis Optica Spectrum Disorder^32^, has identified several polymorphisms in the FCGR2A gene linked to impairment of the mechanism responsible for anti-CD20-mediated B-cell depletion and reduced treatment benefits^31^.

This pilot study has limitations, including that it was from a single center with a limited number of patients studied. In this context, traditional methods to evaluate false discovery rates (FDR) may at times be overly conservative, leading to Type II errors (i.e., false negatives). Therefore, we utilized a multi-step approach that considered multiple statistical methods, and only defined as SNPs of interest those that met at least two of these criteria of significance (Supplemental Figure 2). The current evidence of overrepresentation in affected AA patients of SNPs of interest will require independent validation in a larger cohort of patients with anti-CD20 therapy across multiple institutions.

Replenishment with early B cell subsets, which are naïve to self-antigens, is rationalized as associated with a good short-term prognosis as these subsets of B cells are not known to have pathogenic roles in MS^34-38^. However, there are several caveats to this finding: the number of ER AA MS patients was small; the period of therapy was about 5 years on average, which does not allow us to comment on longer-term outcomes. Recent studies, in which the time between anti-CD20 infusions was extended, have not translated into higher rates of MS lesions on MRI or clinical relapses^34-38^. Yet, ‘progression independent of relapses and MRI lesions’ (PIRMA) may be the main driver of disease progression in MS^39^. Whether patients will manifest similar low relapse and disability outcomes over the longer term, especially the potentially at-risk AA patients with the biology of the ER group that we have identified in our investigations, warrants close investigation^39^.

Despite the higher incidence of MS among female AA patients than other racial-ethnic groups^9^, relatively few studies have focused on the clinical, immunologic, or pathologic features of MS in AA patients. Our study on MS patients of African descent was made possible by the large cohort of AA patients with MS who receive their care at NYU MS Comprehensive Care Center – 20% of the individuals in the MS center self-identify as being AA. Social and economic factors are common confounders in studies of under-represented groups and likely contribute to worse outcomes seen in AA patients with MS^41^. Our participants receiving care in a specialized tertiary center with uniform treatment protocols, reduce but cannot eliminate such concerns. While acknowledging the importance of socio-economic factors, our analyses suggest that immunogenetic factors are may be contributory drivers in some patients.

Our study highlight gaps in the literature as there haven’t been pharmacogenetic studies specifically investigating the relationship between genetic variations and biological, clinical, and radiographic outcomes of anti-CD20 therapy in MS patients^31^. There is interest that higher in vivo anti-CD20 antibody levels and deeper B cell depletion may convey greater clinical benefits (i.e. lower risk of progression independent of relapses)^40^, and here we have a biologic basis for concern regarding treatment efficacy in a subset of AA early repleters. Given the expanding number of anti-CD20 agents approved for MS, future pharmacogenetic studies in larger cohorts that integrate studies of SNPs affecting ADCC, and B cell survival could help predict and optimize therapeutic outcomes. Our study also raises an important question of whether differences in B cell repopulation kinetics in individuals of African descent could contribute to differences in clinical disease course in AA patients with MS.

In conclusion, our study in AA patients identifies that a B cell extrinsic factor, the induction of anti-drug antibodies, as responsible for earlier B-cell repopulation in only small minority of the patients, while in other AA patients we identified a set of SNPs with significant overrepresentation of pathways related to inflammatory response, leukocyte activation, and B cell activation and differentiation. These SNPs could potentially be used for timing of CD20-depleting therapy based on precision medicine considerations and may be potentially relevant to define the risk of disease progression. As there is now a trend in clinical practice to extend the interval between dosing to minimize infectious complications in anti-CD20 treated MS patients, it is important to understand how this strategy will affect patients with rapid repletion, including AA patients who appear to be disproportionately overrepresented.

## Data Availability

All data produced in the present study are available upon reasonable request to the authors

## Acknowledgements

Flow cytometry was performed by the Immune Monitoring Laboratory (RRID: SCR_017936) Division of Advanced Research Technologies, NYU Grossman School of Medicine. This shared resource is partially supported by the NYU Cancer Center Support Grant P30CA016087 at the Laura and Isaac Perlmutter Cancer Center (PCC). ANA was supported in part by NIH 5T32AR069515-07. We thank all our patients for their participation in this study. GJS is supported in part by R01-AR42455 and R21AI180737. We are grateful to Carolyn Akers and Madeline Xin for their help in preparing a supplemental figure. Special thanks to the Parekh Center for Interdisciplinary Neurology at NYU School of Medicine for supporting our work.

## Author Contributions

Study design: GJS and IK. Analysis of genetic data: DF and ANA. Flow cytometric analysis: ANA and AAA. Figure formulation: ANA. Manuscript writing GJS, ANA, and IK. Manuscript editing GJS, ANA, IK, DF, HK, AK, and AAA. GJS had access to all data and attests to data integrity.

---

**Supp Figure 1.**
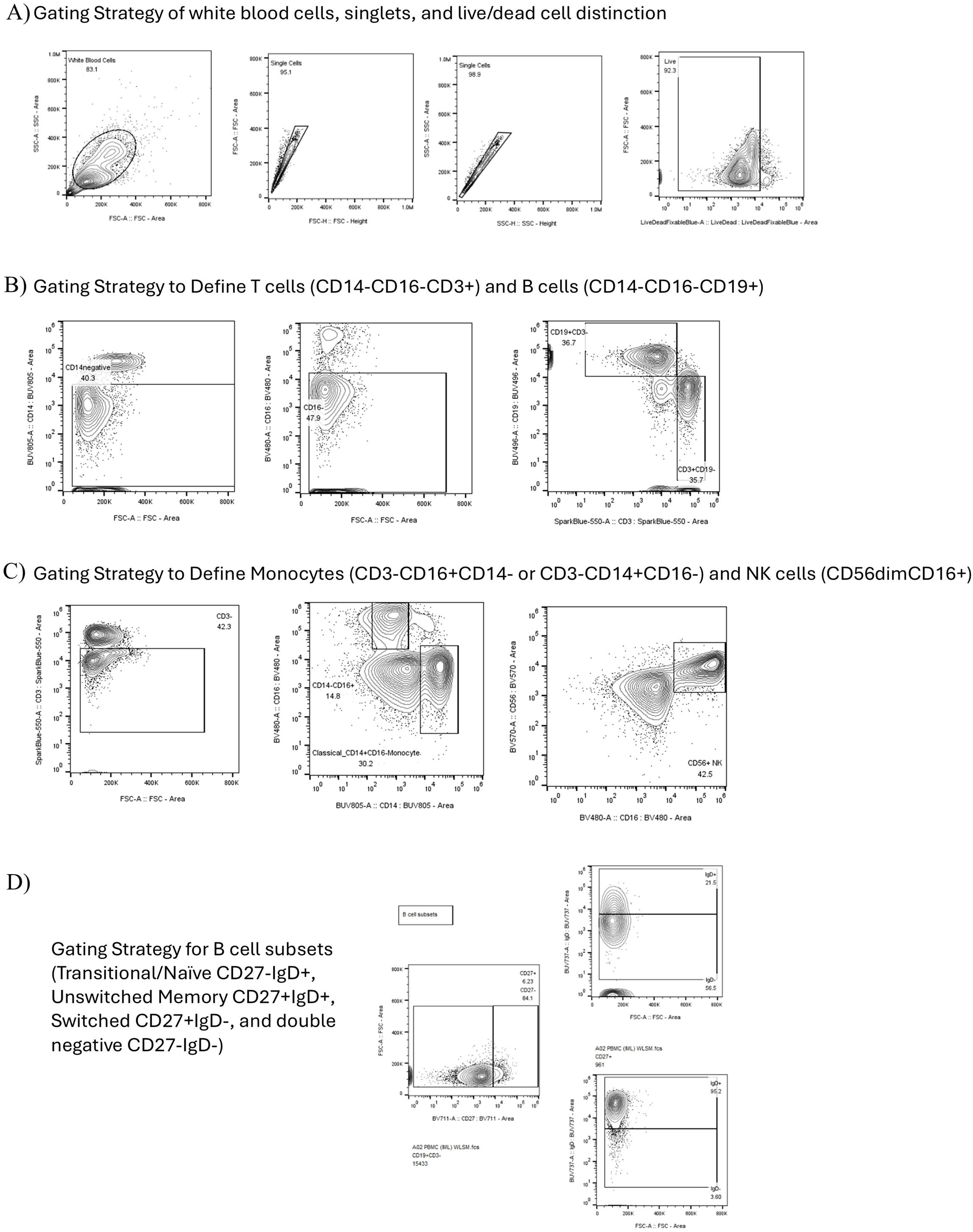

**Supp Figure 1 (E-F).**
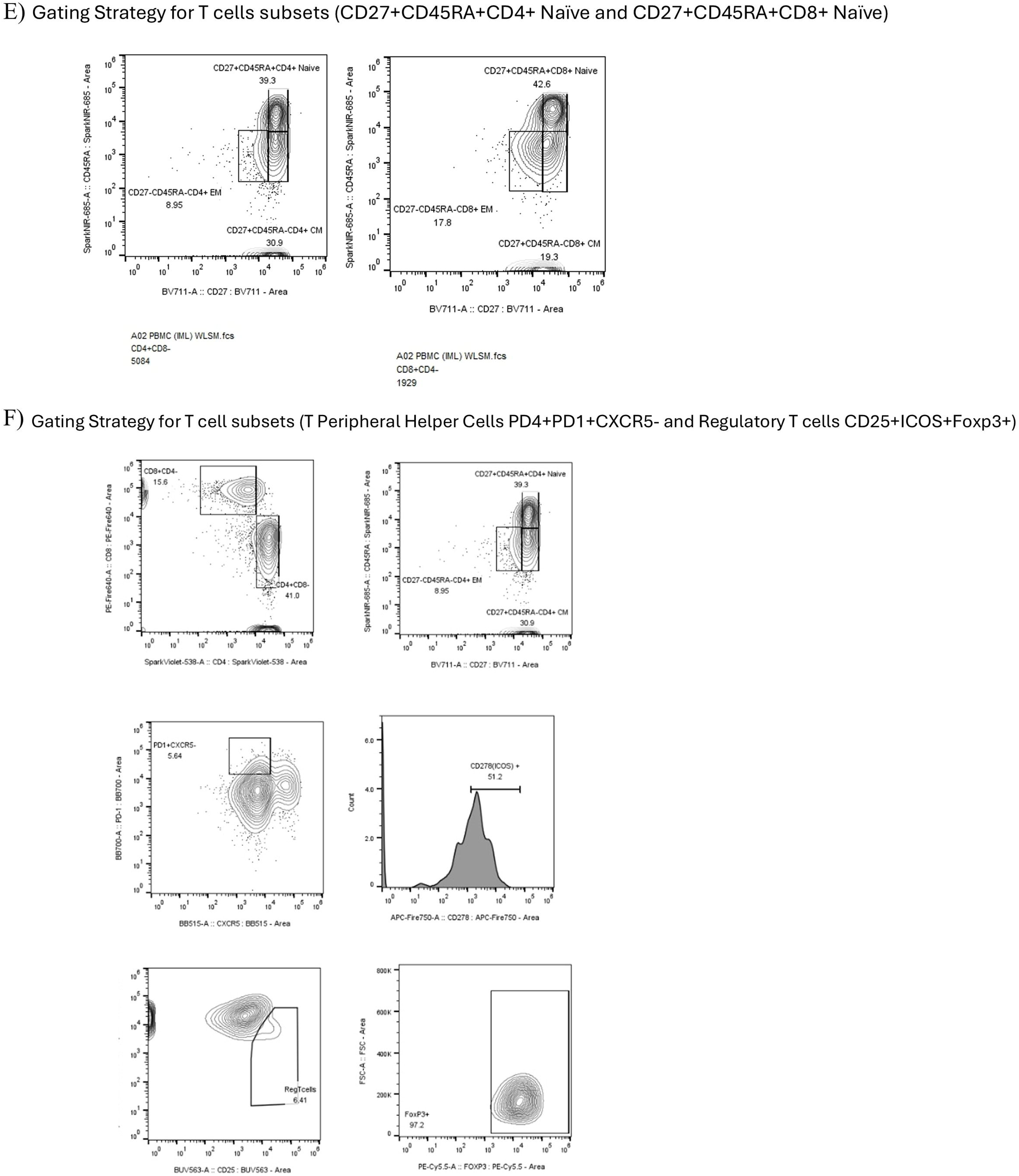

**Supp Figure 2.**
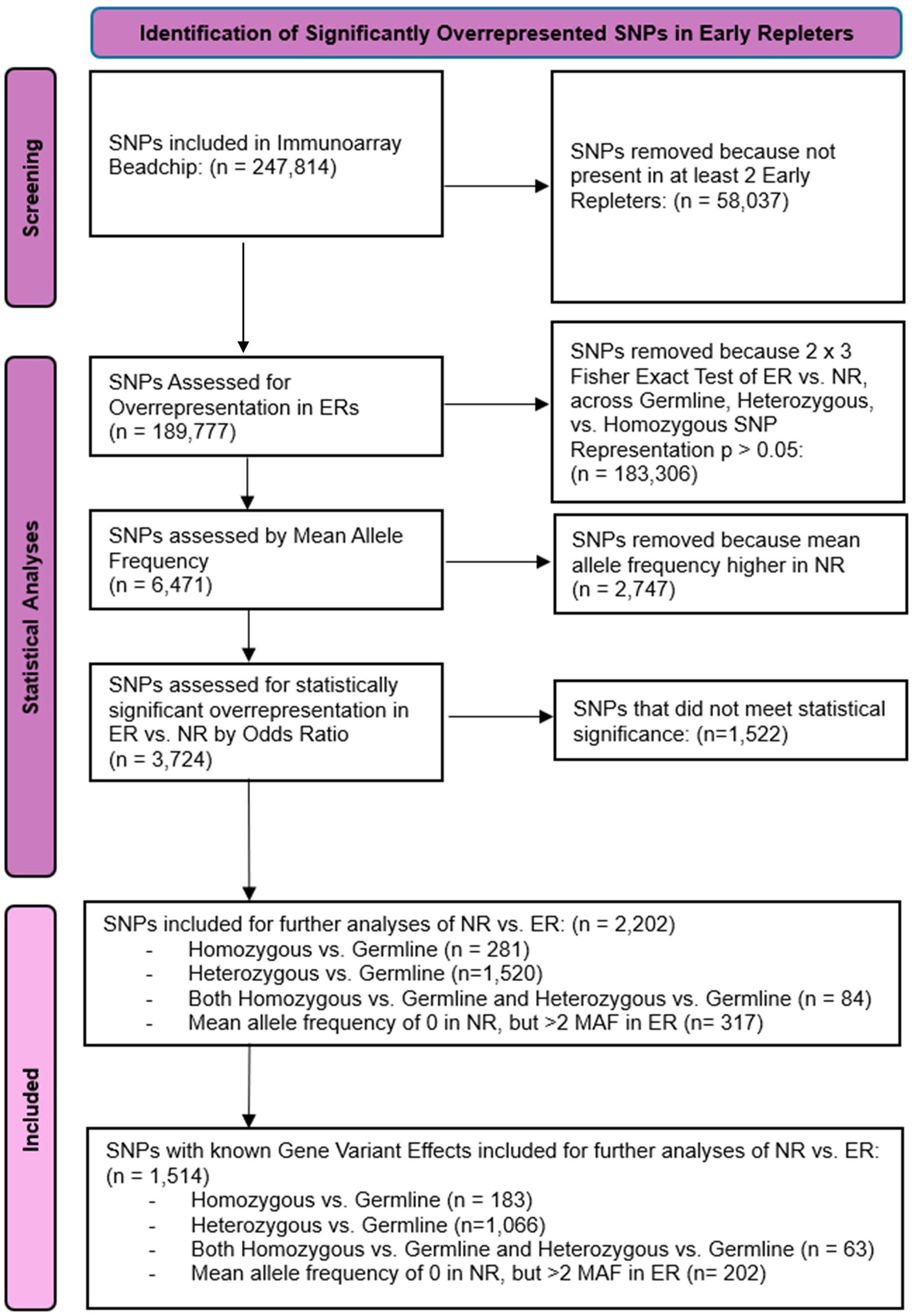

**Supp Figure 3.**
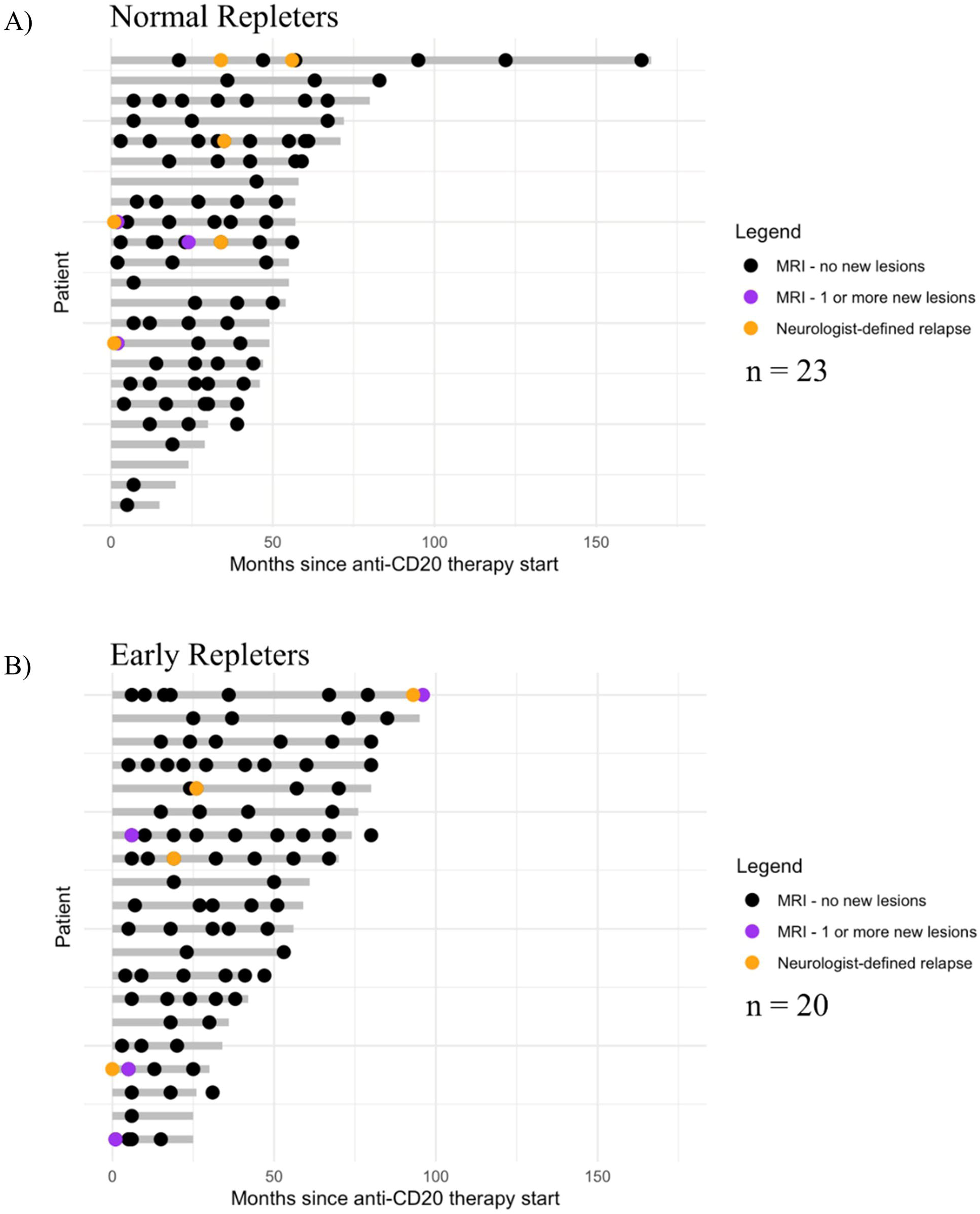

**Supp figure 4.**
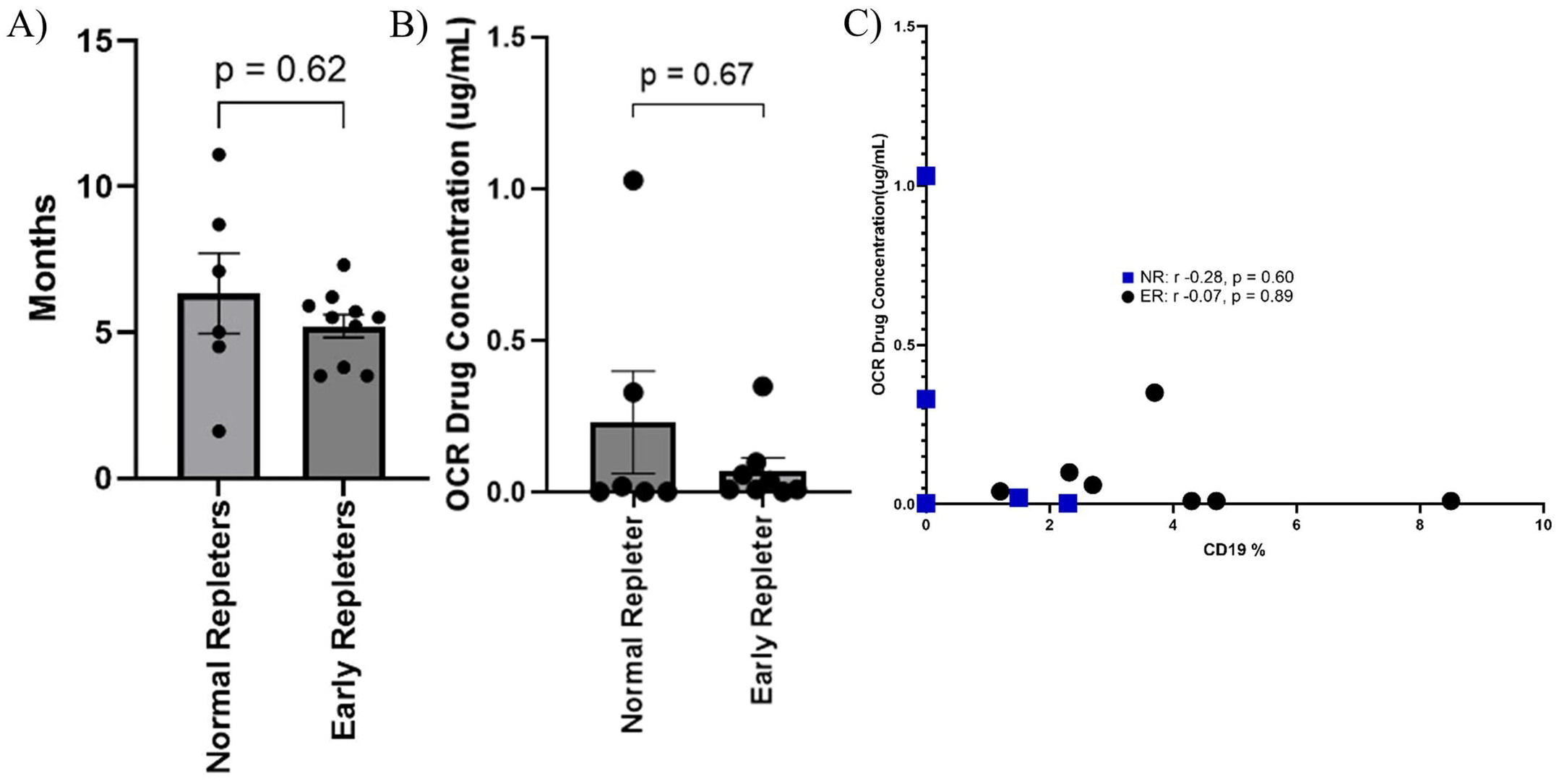

**Supp figure 5 A-F.**
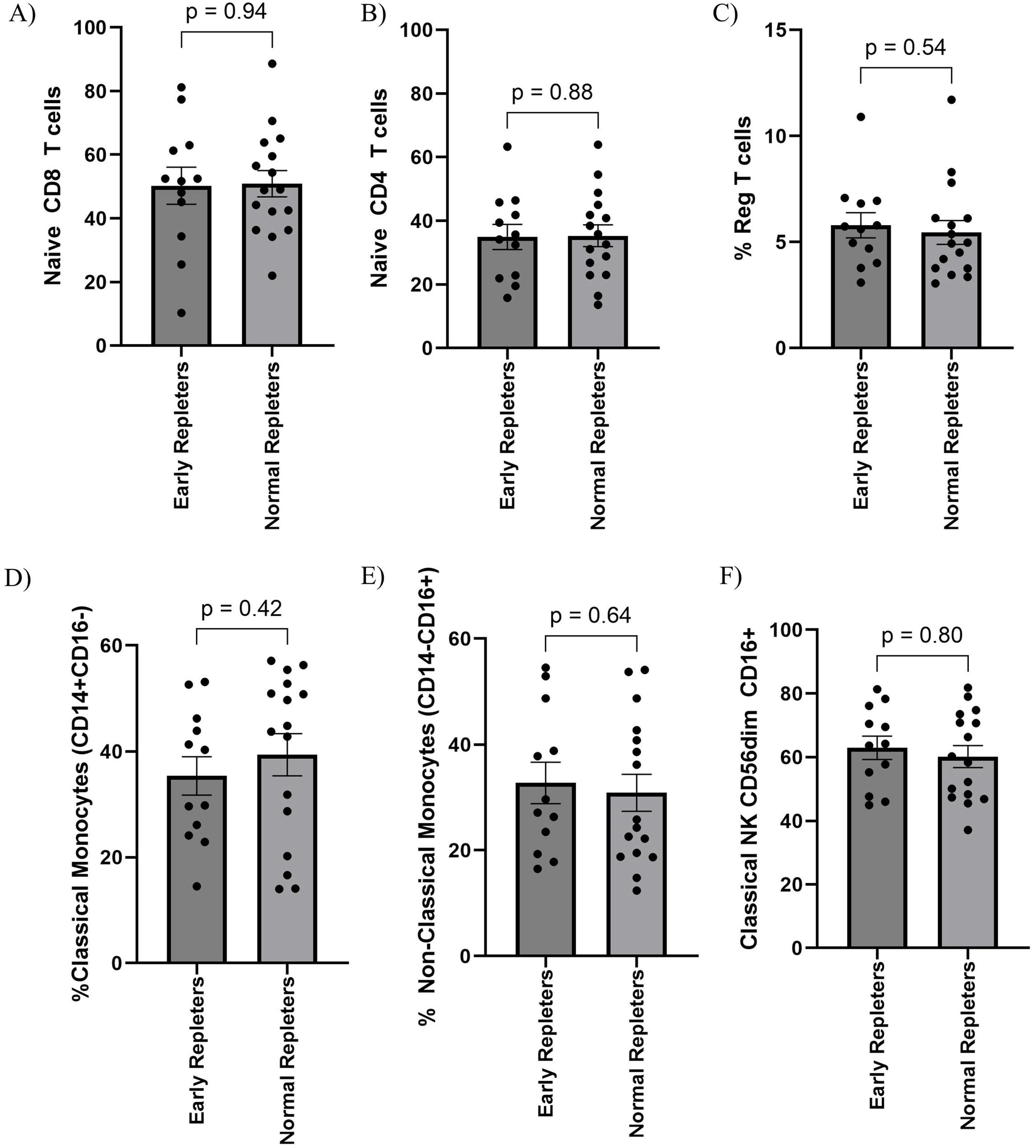

**Supp figure 5 G-K.**
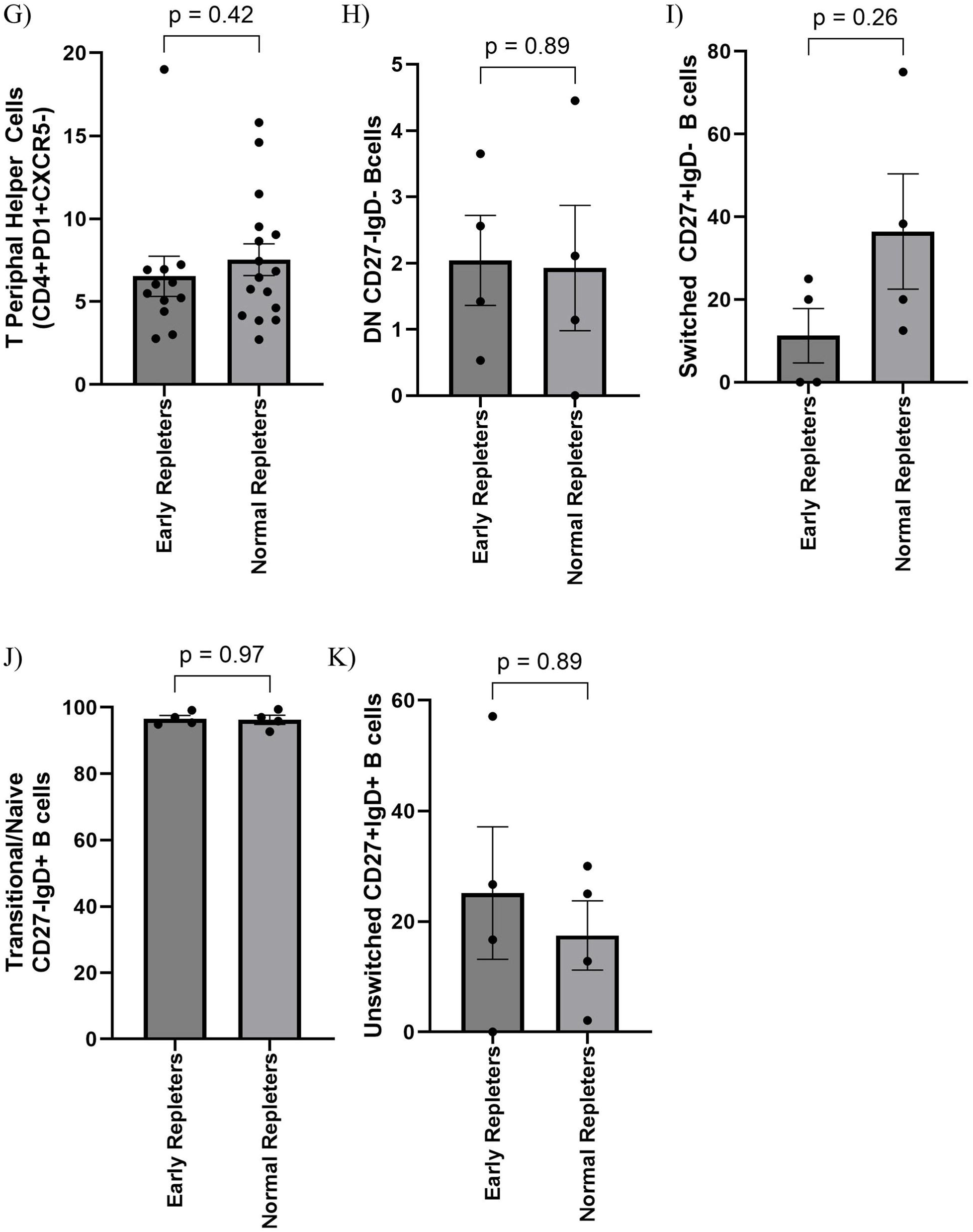

**Supp figure 6.**
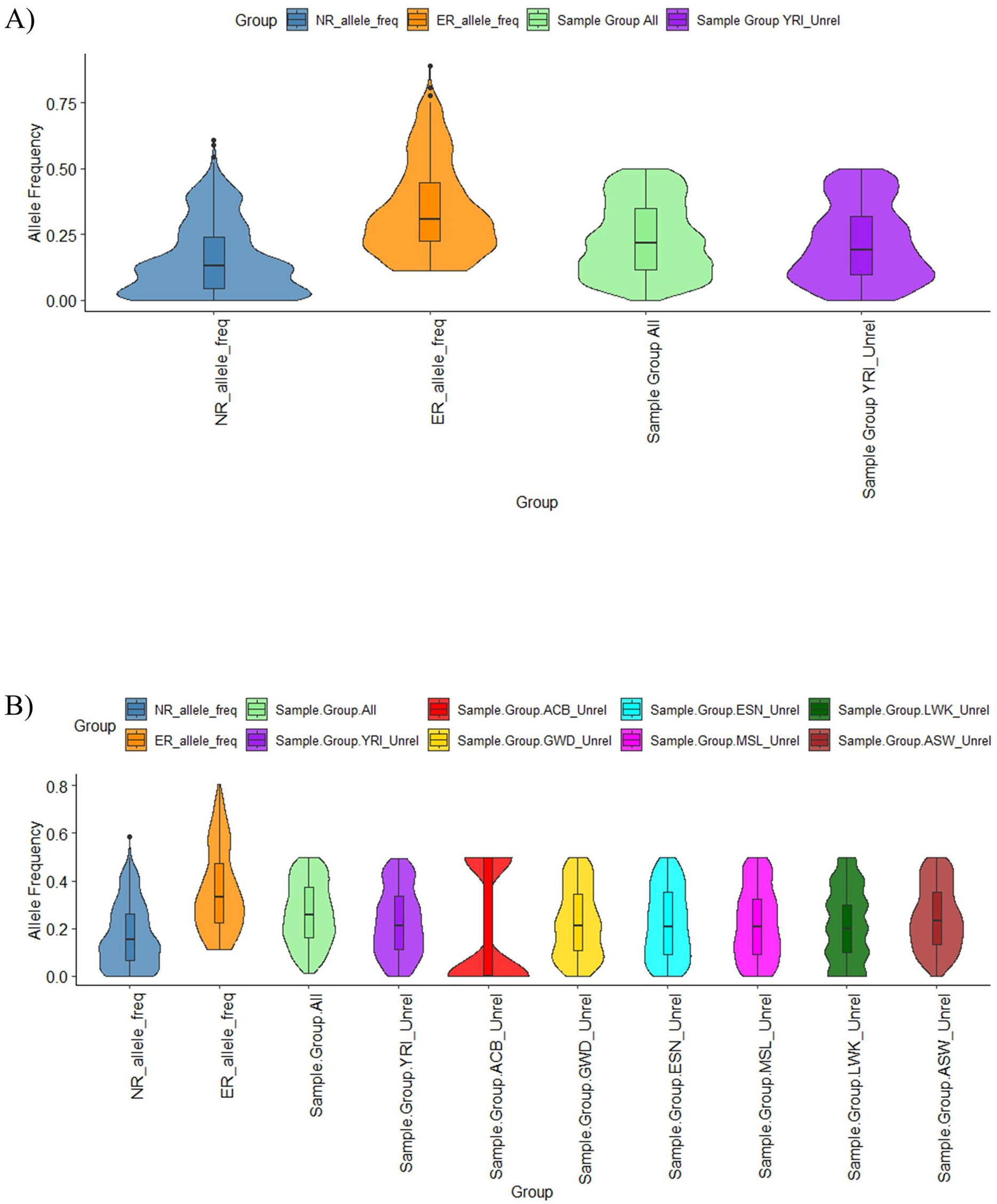

## References

1 Baecher-Allan, C., Kaskow, B. J. & Weiner, H. L. Multiple sclerosis: mechanisms and immunotherapy. Neuron 97, 742–768 (2018).

2 Li, R., Patterson, K. R. & Bar-Or, A. Reassessing B cell contributions in multiple sclerosis. Nature immunology 19, 696–707 (2018).

3 Reich, D. S., Lucchinetti, C. F. & Calabresi, P. A. Multiple Sclerosis. N Engl J Med 378, 169–180, doi:10.1056/NEJMra1401483 (2018).

4 Cantoni, C. et al. A single-cell compendium of human cerebrospinal fluid identifies disease-associated immune cell populations. The Journal of Clinical Investigation 135 (2025).

5 Fernandez Zapata, C., et al. Comprehensive cerebrospinal fluid analysis indicates key roles for B cells in multiple sclerosis. medRxiv, 2025.2001. 2002.24319302 (2025).

6 Hauser, S. L. et al. Ocrelizumab versus interferon beta-1a in relapsing multiple sclerosis. New England Journal of Medicine 376, 221–234 (2017).

7 Margoni, M., Preziosa, P., Filippi, M. & Rocca, M. A. Anti-CD20 therapies for multiple sclerosis: current status and future perspectives. Journal of Neurology 269, 1316–1334 (2022).

8 Hittle, M. et al. Population-based estimates for the prevalence of multiple sclerosis in the United States by race, ethnicity, age, sex, and geographic region. JAMA neurology 80, 693–701 (2023).

9 Langer-Gould, A., Brara, S. M., Beaber, B. E. & Zhang, J. L. Incidence of multiple sclerosis in multiple racial and ethnic groups. Neurology 80, 1734–1739, doi:10.1212/WNL.0b013e3182918cc2 (2013).

10 Khan, O. et al. Multiple sclerosis in US minority populations: clinical practice insights. Neurology: Clinical Practice 5, 132–142 (2015).

11 Saidenberg, L. et al. Faster B-cell repletion after anti-CD20 infusion in Black patients compared to white patients with neurologic diseases. 63, 103830 (2022).

12 Thompson, A. J. et al. Diagnosis of multiple sclerosis: 2017 revisions of the McDonald criteria. The Lancet Neurology 17, 162–173 (2018).

13 Kolberg, L. et al. g: Profiler—interoperable web service for functional enrichment analysis and gene identifier mapping (2023 update). Nucleic acids research 51, W207–W212 (2023).

14 Kassambara, A. et al. RNA-sequencing data-driven dissection of human plasma cell diferentiation reveals new potential transcription regulators. 35, 1451–1462 (2021).

15 Kassambara, A. et al. GenomicScape: an easy-to-use web tool for gene expression data analysis. Application to investigate the molecular events in the diferentiation of B cells into plasma cells. PLoS computational biology 11, e1004077 (2015).

16 Lee, R. D. et al. Single-cell analysis identifies dynamic gene expression networks that govern B cell development and transformation. Nature communications 12, 6843 (2021).

17 Salerno, F. et al. An integrated proteome and transcriptome of B cell maturation defines poised activation states of transitional and mature B cells. Nature Communications 14, 5116 (2023).

18 Ntellas, P. et al. TNFRSF13C/BAFFR P21R and H159Y polymorphisms in multiple sclerosis. Multiple Sclerosis and Related Disorders 37, 101422 (2020).

19 Kornilov, S. A. et al. Multi-Omic characterization of the efects of ocrelizumab in patients with relapsing-remitting multiple sclerosis. Journal of the Neurological Sciences 467, 123303 (2024).

20 Itoh-Nakadai, A. et al. The transcription repressors Bach2 and Bach1 promote B cell development by repressing the myeloid program. Nature immunology 15, 1171–1180 (2014).

21 Isobe, N. et al. An ImmunoChip study of multiple sclerosis risk in African Americans. 138, 1518–1530 (2015).

22 Xue, H., Arbini, A. A., Melton, H. J., Kister, I. J. M. S. & Disorders, R. African American patients with Multiple Sclerosis (MS) have higher proportions of CD19+ and CD20+ B-cell lineage cells in their cerebrospinal fluid than White MS patients. 79, 105047 (2023).

23 Telesford, K. M. et al. Understanding humoral immunity and multiple sclerosis severity in Black, and Latinx patients. 14, 1172993 (2023).

24 da Gama, P. D. et al. Oligoclonal Bands in Cerebrospinal Fluid of Black Patients with Multiple Sclerosis. Biomed Res Int 2015, 217961, doi:10.1155/2015/217961 (2015).

25 Menard, L. C. et al. B cells from African American lupus patients exhibit an activated phenotype. JCI Insight 1, e87310, doi:10.1172/jci.insight.87310 (2016).

26 Steri, M. et al. Overexpression of the Cytokine BAFF and Autoimmunity Risk. N Engl J Med 376, 1615–1626, doi:10.1056/NEJMoa1610528 (2017).

27 Ritterhouse, L. L. et al. B lymphocyte stimulator levels in systemic lupus erythematosus: higher circulating levels in African American patients and increased production after influenza vaccination in patients with low baseline levels. Arthritis & Rheumatism 63, 3931–3941 (2011).

28 Tollerud, D. J. et al. The influence of age, race, and gender on peripheral blood mononuclear-cell subsets in healthy nonsmokers. J Clin Immunol 9, 214–222, doi:10.1007/BF00916817 (1989).

29 Kurupati, R. et al. Race-related diferences in antibody responses to the inactivated influenza vaccine are linked to distinct pre-vaccination gene expression profiles in blood. Oncotarget 7, 62898–62911, doi:10.18632/oncotarget.11704 (2016).

30 Wang, S. S. et al. Common gene variants in the tumor necrosis factor (TNF) and TNF receptor superfamilies and NF-kB transcription factors and non-Hodgkin lymphoma risk. PloS one 4, e5360 (2009).

31 Zhong, M. et al. The pharmacogenetics of rituximab: Potential implications for anti-CD20 therapies in multiple sclerosis. Neurotherapeutics 17, 1768–1784 (2020).

32 Kim, S.-H. et al. Treatment outcomes with rituximab in 100 patients with neuromyelitis optica: influence of FCGR3A polymorphisms on the therapeutic response to rituximab. JAMA neurology 72, 989–995 (2015).

33 Novak, A. J. et al. Elevated serum B-lymphocyte stimulator levels in patients with familial lymphoproliferative disorders. Journal of clinical oncology 24, 983–987 (2006).

34 Hogenboom, L., van Kempen, Z. L., Kalincik, T., Bar-Or, A. & Killestein, J. A personalized approach for anti-CD20 therapies in multiple sclerosis. Multiple Sclerosis and Related Disorders 91, 105851 (2024).

35 Rempe, T. et al. Ocrelizumab B-cell repopulation-guided extended interval dosing versus standard dosing–similar clinical eficacy with decreased immunoglobulin M deficiency rates. Multiple Sclerosis and Related Disorders 79, 105028 (2023).

36 Bou Rjeily, N., Fitzgerald, K. C. & Mowry, E. M. Extended interval dosing of ocrelizumab in patients with multiple sclerosis is not associated with meaningful diferences in disease activity. Multiple Sclerosis Journal 30, 257–260 (2024).

37 Rodriguez-Mogeda, C. et al. Extended interval dosing of ocrelizumab modifies the repopulation of B cells without altering the clinical eficacy in multiple sclerosis. Journal of neuroinflammation 20, 215 (2023).

38 Baig, M. M. A. et al. Comparing the eficacy and safety of extended vs standard dosing of ocrelizumab in MS: A systemic review and meta-analysis. Multiple Sclerosis and Related Disorders, 106257 (2025).

39 Kapica-Topczewska, K. et al. Assessment of disability progression independent of relapse and brain MRI activity in patients with multiple sclerosis in Poland. Journal of Clinical Medicine 10, 868 (2021).

40 Hauser, S. L. et al. Association of higher ocrelizumab exposure with reduced disability progression in multiple sclerosis. Neurology: Neuroimmunology & Neuroinflammation 10, e200094 (2023).

41 Brayo, P. & Kimbrough, D. Multiple sclerosis in the Black population of the United States. Practical Neurology [Internet] (2021).

