## Supplementary figures and images for "B cell-extrinsic and intrinsic factors linked to early immune repletion after anti-CD20 therapy in patients with Multiple Sclerosis of African Ancestry"

### Supplemental Table 1

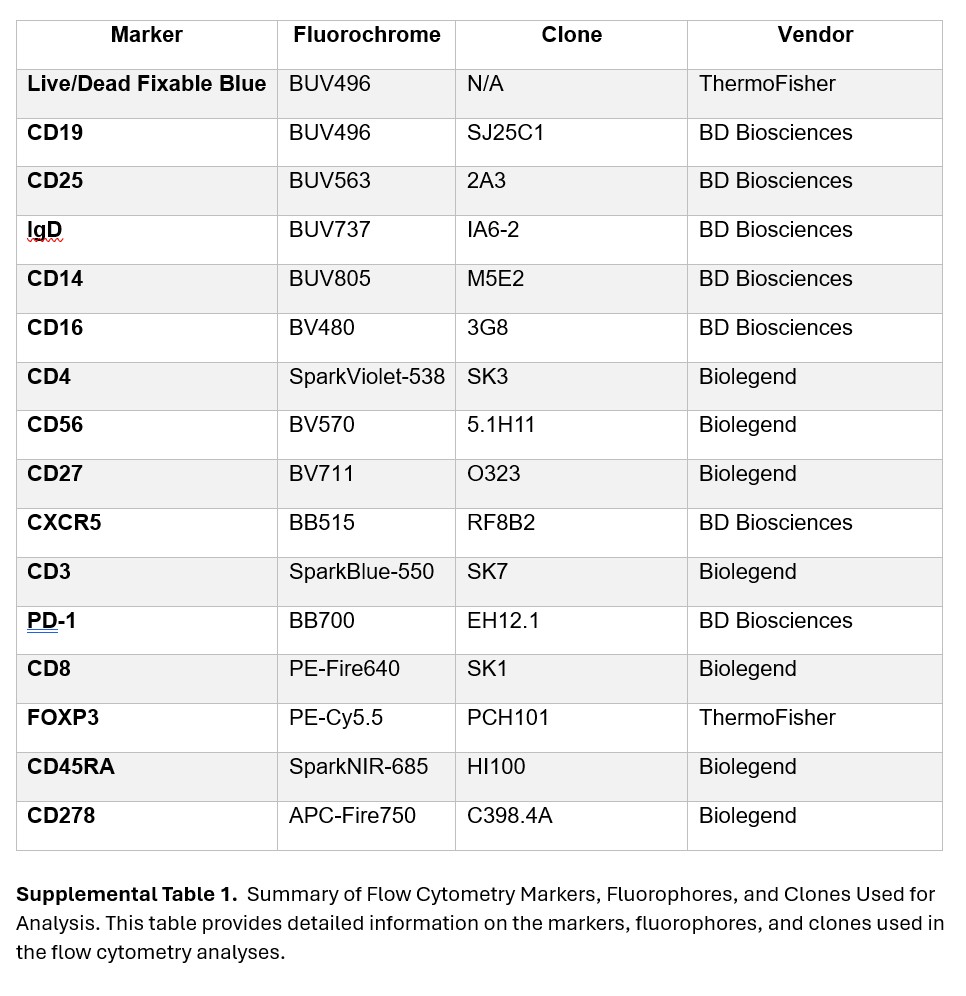
