## Supplemental Table and Figure Legends for "B cell-extrinsic and intrinsic factors linked to early immune repletion after anti-CD20 therapy in patients with Multiple Sclerosis of African Ancestry"

Supplemental Legends and Tables

Legends for Supplemental Table 2,3,4 and 5 (All .xls files)

**Supplemental Table 2.** 2,202 Genes that are Overrepresented in Early Repleters (ER) compared to Normal Repleters (NR). The table provides a comprehensive list of genes that are significantly overrepresented in ERs. This table includes the following columns: Gene Symbol, Gene Name, Chr (chromosome), Start Position, End Position, Number of SNPs, and gene variant effects.

**Supplemental Table 3**. Summary of Pathway Analysis Genes and Overlapping Genes. This table highlights the genes identified through pathway analysis that are significantly associated with the condition of interest and their overlap with other relevant gene sets.

**Supplemental Table 4.** Summary of SNPs overrepresented in patients with high sCD40L compared to other early repleters (ER). Highlights the genetic variations that are significantly overrepresented in patients with high serum volumes of sCD40L compared to other ERs. Variant effects are described, defined by the Sequence Ontology, and ordered by severity from g:SNPense.

**Supplemental Table 5.** Summary of SNPs overrepresented in superrepleter patients compared to other early repleters (ER). Highlights the genetic variations that are significantly overrepresented in superrepleter patients. Variant effects are described, defined by the Sequence Ontology, and ordered by severity from g:SNPense.

Legends for Supplemental Figures

**Supplemental Figure 1.** Gating strategy for Flow Cytometry Studies. Flow cytometry gating strategy used to identify and analyze B cells and various B cell subsets. The gating strategy includes the initial identification of lymphocytes followed by the isolation of CD19+ B cells. Subsequent gates are applied to distinguish between naive B cells, memory B cells, activated naive B cells, and DN2 B cells. Each gating step is shown with representative flow cytometry plots.

**Supplemental Figure 2.** Flow diagram illustrating the identification of significantly overrepresented SNPs in early repleters (ERs). The diagram outlines the process starting from the initial screen of SNPs included in the study, followed by multiple statistical assessments to refine the list of SNPs.

**Supplemental Figure 3.** Clinical and MRI activity in normal repleters (NR) and early repleters (ER). Number of MRI evaluations completed per patient assessed (black), with timeline of new MRI lesions (purple) and neurologist-defined relapses (orange).

**Supplemental Figure 4.** Correlation of OCR drug concentration and CD19% B cell percentages. (Left) No difference in time since infusion for samples collected for assessment of anti-drug antibodies or ocrelizumab drug concentrations. (Middle) No difference in overall drug concentration between NR (normal repleters) and ER (early repleters) patients studied (n = 6 NRs, n = 8 ERs). (Right) Spearman correlation of CD19% and OCR drug concentration.

**Supplemental Figure 5.** Summary of the proportions of immune cell subsets in expression ER and NR participantsratio (ER) and non-responders (NR) patients. The following B cell subsets were assessed: transitional/naïve (CD27⁻IgD⁺), switched memory (CD27⁺IgD⁻), unswitched memory (CD27⁺IgD⁺), and double negative (CD27⁻IgD⁻) B cells in multiple sclerosis (MS) patients versus non-responders (NRs). Additionally, no significant differences were observed in the peripheral blood representation of naïve CD4⁺ T cells, naïve CD8⁺ T cells, regulatory T cells (CD4⁺CD25⁺), or T peripheral helper cells (TPH) (CD4⁺PD1⁺CXCR5⁻). Mean and standard error of the mean (SEM) for each subset are shown. For monocytes, NK cells, and T cells, samples for flow cytometry were tested at 4.7 ± .83 months since infusion for ER and 5.09 ± 2.3 months for NR. For B cells, only flow cytometry samples where a minimum of CD19⁺ B cells were available for subset analysis were included. For B cell subset analyses, the time since infusion was 5.2 ± .6 months for ER and 7.15 ± 4. months for NR. Markers were not available to differentiate transitional versus naïve B cell subsets further.

**Supplemental Figure 6.** (A) Mean allele frequency (MAF) distribution of SNPs of interest (2171 SNPs) in normal repleter (n = 23) and early repleter (n = 18) patients, compared to reference MAF of unrelated individuals. From data in the public domain, the reference populations include the Yoruba people from Ibadan, Nigeria (YRI; n = 58), and a diverse sample group of unrelated individuals (Sample Group All) from the Yoruba in Ibadan, Nigeria (YRI), Han Chinese in Beijing, China (CHB), Japanese in Tokyo, Japan (JPT), and Utah residents with Northern and Western European ancestry (CEU), totaling n = 201. The YRI and all sample reference group MAFs are sourced from the Illumina ImmunoArray Population report. (B) Mean allele frequency (MAF) distribution of SNPs of interest that overlap with reference SNP mean allele frequency data from the Illumina global diversity array (475 SNPs) in normal repleter (n = 23) and early repleter (n = 18) patients compared to Supplemental Figure 6. Mean allele frequency (MAF) distribution of SNPs of interest that overlap with reference SNP mean allele frequency data from the Illumina global diversity array (475 SNPs) in normal repleter (n = 23) and early repleter (n = 18) patients, compared to reference MAF of unrelated individuals. The reference populations include a diverse sample group of unrelated individuals, totaling n = 2051. The specific reference populations and their sample sizes are as follows: Yoruba people from Ibadan, Nigeria (YRI; n = 85), African Caribbeans in Barbados (ACB; n = 1), Gambian in Western Divisions in the Gambia (GWD; n = 19), Esan in Nigeria (ESN; n = 17), Mende in Sierra Leone (MSL; n = 17), Luhya in Webuye, Kenya (LWK; n = 5), and Americans of African Ancestry in SW USA (ASW; n = 56). The MAFs for these reference populations are sourced from the Illumina global diversity array reference report.
